# Digital biomarkers for insulin resistance screening in daily life

**DOI:** 10.64898/2026.05.20.26353669

**Authors:** Mia Jovanova, Victoria Bruegger, Radoslava Švihrová, Magdalena Fuchs, Qiuhan Jin, Felix Wortmann, Michael Mitter, Michal Bechny

**Author notes:** Corresponding author: Dr. Mia Jovanova, Dufourstrasse 40a, 9000 St. Gallen, Switzerland.

## Abstract

One in four adults has insulin resistance (IR), a modifiable driver of type-2 diabetes that can precede diagnosis by a decade. However, IR assessment remains clinic- and laboratory-based, limiting repeated population screening. We tested whether free-living wearable data can detect IR in adults with normoglycemia or prediabetes. Machine-learning models using continuous glucose monitor(CGM)-based glucose dynamics and smartwatch-based heart rate/heart rate variability were developed in Study 1 (*N* = 97) and externally validated without retraining in Study 2 (*N* = 61, 31% IR prevalence). The best-performing CGM-based model achieved AU-ROC = 0.873 [0.756–0.967] and AU-PRC = 0.816 [0.640–0.934], outperforming an anthropometrics-only baseline (AU-ROC = 0.749, AU-PRC = 0.593). Findings are the first to detect IR from wearables without blood tests or structured glucose challenges, with state-of-the-art comparable performance. By enabling continuous at-home screening, this approach can identify undetected at-risk individuals and trigger confirmatory blood tests to close detection gaps.

## Introduction

Type-2 diabetes (T2D) represents one of the most prevalent and economically consequential chronic diseases globally^1^, affecting an estimated 589 million adults, a figure projected to rise to 853 million by 2050^2^. Approximately 90% of all cases are type-2 diabetes^3^, a disease strongly influenced by modifiable lifestyle factors such as diet, physical activity, and weight management^4–6^. Direct healthcare expenditure attributable to diabetes—predominantly through complications that are, in large part, preventable, exceeded USD $1 trillion globally in 2024^7^, representing a 338% increase over the preceding 17 years. When indirect costs are considered, the macroeconomic burden is estimated at $10.2 trillion between 2020 and 2050^2,7^. Overall, diabetes consumes nearly one in eight dollars of global health spending, underscoring an urgent need for earlier risk detection and intervention at scale.

Central to the type-2 diabetes disease trajectory is insulin resistance (IR), a key precursor state in which skeletal muscle, adipose, and liver cells become less responsive to insulin. IR reduces glucose uptake from the blood and drives compensatory increases in insulin secretion by the pancreas, which over time lead to beta-cell failure, rising blood glucose levels, and chronic hyperglycaemia^8–10^. IR often precedes clinical type-2 diabetes diagnosis by ten to 15 years, constituting an early target state in the disease continuum^10–13^. Critically, early IR is modifiable: lifestyle-induced weight loss interventions can improve insulin sensitivity in individuals with impaired glucose tolerance^14^. Lifestyle interventions are 44% more likely to reverse prediabetes to normoglycaemia (when compared with usual care, i.e., standard physician advice), based on a recent meta-analysis of 31 randomized control trials enrolling 23,684 adults with prediabetes^15^. Specifically, the Diabetes Prevention Program, Finnish Diabetes Prevention Study, and Da Qing trials, collectively show a 30–60% reduction in type-2 diabetes incidence, through structured dietary and physical activity modification initiated at an early stage of impaired glucose tolerance, prior to overt hyperglycaemia^4,5,16^. Critically, the effectiveness of such lifestyle interventions is largely contingent on early risk identification.

In contrast, detection of IR remains largely absent from routine clinical practice. The diagnostic standard—the hyperinsulinaemic–euglycaemic clamp, requires intravenous insulin and glucose infusion with repeated arterialised blood sampling over six or more hours under controlled clinical conditions, precluding deployment in routine or community-based care^17^. Established surrogates, such as the Homeostatic Model Assessment of Insulin Resistance (HOMA-IR), estimate insulin sensitivity from fasting glucose and fasting insulin via venepuncture, followed by centralised laboratory processing^18,19^.While cheaper than the hyperinsulinaemic–euglycaemic clamp, HOMA-IR’s in still dependent on standardized insulin assays and access to laboratory infrastructure. Relatedly, an estimated 252 million adults are expected to live undiagnosed diabetes, predominantly type-2 diabetes, while a further 635 million live with undetected impaired glucose tolerance^20^. At this scale, the absence of scalable, population-level screening for IR, a key early pathophysiological marker of metabolic dysfunction, limits opportunities for timely prevention. Critically, the absence of population IR screening allows the window of modifiable metabolic risk to pass largely undetected.

Recent advances in consumer-grade wearable technology present new opportunities for population-level IR screening outside traditional clinical infrastructure, initially in digitally engaged populations, and in developed countries. There are approximately 560 million smartwatch users worldwide, representing over a quarter of adults globally in developed countries^21,22^, with the majority using their devices primarily for health and fitness monitoring. Continuous glucose monitors (CGM), which sample interstitial glucose at approximately five-minute intervals, are undergoing a parallel transition. In 2024, the US Food and Drug Administration approved over-the-counter CGMs for individuals without diabetes^23^, and their use in non-diabetic populations for lifestyle optimization is expanding^24,25^. Together, these two technologies present plausible IR screening pathways that could be deployable at home, entirely outside clinical settings.

How might consumer smartwatches and CGMs help track IR in non-clinical settings? Physiologically, IR is associated with autonomic dysregulation, namely attenuated parasympathetic activity and elevated sympathetic tone; manifesting as elevated resting heart rate (HR) and reduced heart rate variability (HRV)^26,27^, which is detectable via consumer smartwatches^28^. In normoglycemic individuals, heart rate naturally drops at night as the body shifts into a restorative parasympathetic state. In individuals with IR, persistent sympathetic overactivation may blunt this nocturnal recovery^29^, potentially resulting in greater day-to-day variability in the day-to-night heart rate transition^30^. In parallel, IR impairs glucose homeostasis by reducing the body’s ability to efficiently clear glucose from the bloodstream^9^. Compared with insulin-sensitive individuals, those with IR show higher glucose peaks, broader postprandial curves, and slower return to baseline^31^; as well as greater glucose variability under free-living conditions^32^. IR is also expected to correlate with higher fasting glucose in the wake-to-first-meal window, due to impaired overnight suppression of hepatic glucose production^9^, although this has not yet been directly demonstrated in free-living CGM studies of non-diabetic populations. Together, these haemodynamic and glycemic signatures, detectable via smartwatches and CGMs, offer a biologically grounded basis for detecting IR continuously in free-living settings.

Recent studies have begun to develop AI-based algorithms for wearable-based IR prediction. Published in *Nature Biomedical Engineerin*g, Ref. ^31^ demonstrated that machine-learning models applied to CGM glucose curves from structured at-home oral glucose tolerance tests predict muscle IR with Area Under the Receiver Operating Characteristic Curve (AU-ROC) 0.88, validated against a gold-standard insulin suppression test. However, this approach required a structured glucose challenge rather than passive free-living monitoring and was tested with a small sample (*n* = 29). The recent, seminal WEAR-ME study, published in *Nature*, subsequently trained deep neural networks against HOMA-IR using raw smartwatch time-series combined with routine blood biomarkers in 1,165 participants, achieving AU-ROC 0.80. In an independent validation cohort of 72 participants, a pre-trained wearable foundation model, together with demographics, reached AU-ROC 0.75 — above a demographics-only baseline of 0.66. An AU-ROC 0.88 was only recovered by adding fasting glucose and a full lipid panel, preserving a dependency on venepuncture and centralized laboratory processing^33^. Three critical gaps remain unaddressed. First, no study has evaluated whether passively collected, free-living CGM features, requiring no structured glucose challenge or deliberate participant action, carry independent discriminative value for IR classification, leaving a key modality entirely untested in a passive screening framework. Second, WEAR-ME’s wearable-foundation model achieved a modest improvement over a demographics baseline, and it is unclear whether CGM data could meaningfully augment discrimination. Third, the large-scale pretrained neural network offers limited interpretability, making it difficult to identify which physiological signals drive classification. Model interpretability is a necessary pre-requirement to build clinician trust^34^ required for population-level deployment. No study has examined whether a smaller set of theoretically motivated, pathophysiology-driven features can achieve comparable performance with greater transparency, in a free-living context. Addressing these gaps is an essential first step to establishing whether consumer smartwatches and CGMs could form the basis of a scalable, passive, and interpretable AI-driven IR screening infrastructure; deployable entirely outside clinical settings.

To address these gaps, we developed theory-driven digital biomarkers for IR prediction and evaluated three modality-specific random forest classifiers in adults with normoglycaemia or prediabetes: an anthropometrics-only baseline, a smartwatch-based HR/HRV model, and a passive, free-living CGM model. We used AI-READI^35,36^, a publicly available dataset from three US clinical sites, in which participants underwent a fasting blood draw to establish HOMA-IR ground-truth labels alongside approximately 10 days of concurrent free-living CGM and smartwatch monitoring; in addition to other tests beyond scope. To focus on free-living data collection, no models included laboratory analysis as feature inputs (see Table S1 for full feature list across models). We trained models in the internal cohort (Study 1, *N* = 97) and assessed external generalizability in an independent cohort without model retraining (Study 2, *N* = 61). We used Area under the receiver operating characteristic (AU-ROC) as our primary discrimination metric given its widespread use in digital health prediction (for example, Refs. ^37–39^), and comparability with recent work^33^, alongside the Area Under the Precision–Recall Area (AU-PROC) and Area Under the Precision–Recall Gain Curve (AU-PRGC)^40^, which are better suited to handle class imbalance. We observed an IR prevalence of approximately 32% across both cohorts. Our primary evaluation metrics are derived from Study 2 external validation on an independent, geographically distinct sample. Additional models with wearable-derived physical activity features are presented in Supplement B.

## Results

### Descriptives

Study 1 included 97 participants recruited from two geographically distinct clinical sites (University of Washington and University of California San Diego; with a mean age of 57.9 years (median = 57.0, *SD* = 11.1, 0% missing). Participant eligibility and recruitment is described in ‘Methods: Participants’, the PRISMA flow diagram is provided in Figure S1, and for additional recruitment details see Refs. ^36,41,42^. Thirty-two participants (33.0%) were classified as being IR (HOMA-IR ≥ 2.9). Mean Hemoglobin A1c (HbA1c) was 5.69% (median = 5.70, *SD* = 0.39), mean fasting glucose was 5.32 mmol/L (median = 5.27, *SD* = 0.70), and mean fasting insulin was 88.1 pmol/L (median = 62.0, *SD* = 74.3). On average, participants had 9.6 days of smartwatch data (median = 10.0, *SD* = 2.9; n=95; two participants missing) and 10.8 days of CGM data (median = 11.0, *SD* = 0.8; *n*=96; one participant missing). Average BMI was 30.2 kg/m² (median = 28.1, *SD* = 7.7); average waist circumference was 99.1 cm (median = 96.3, *SD* = 18.0); average hip circumference was 108.7 cm (median = 105.0, *SD* = 16.2); and average waist-to-hip ratio was 0.91 (median = 0.91, *SD* = 0.09).

Study 2 included 61 non-overlapping participants recruited from a single site (University of Alabama), representing a third geographically distinct region of the United States. Participants had a mean age of 56.0 years (median = 56.0, *SD* = 10.2), of whom 31.1% were classified as IR (HOMA-IR ≥ 2.9). Mean HbA1c was 5.63% (median = 5.70, *SD* = 0.45), mean fasting glucose was 4.64 mmol/L (median = 4.61, SD = 0.77), and mean fasting insulin was 97.3 pmol/L (median = 67.2, *SD* = 92.5). On average, participants had 9.0 days of smartwatch data available (median = 10.0, *SD* = 2.6) and *10.7* days of CGM data (median = 11.0, *SD* = 1.4). Average BMI was 33.2 kg/m² (median = 31.2, SD = 9.4). Importantly, our outcome base rate (IR) was similar across both studies (Study 1: 33.0% IR, 32/97; Study 2: 31.1% IR, 19/61; χ²(1) = 0.004, *p* = .947), supporting the feasibility of Study 2 as an external validation sample. Additional descriptive statistics for CGM- and smartwatch-derived variables, by IR status, are presented in Table S2. The distribution of IR status and HbA1c-based glycemic classifications (normoglycemic vs. prediabetic) across both studies is presented in Figure S2 and Table S3. Spearman correlations among key study variables are reported in Table S4.

### Developing Smartwatch and CGM-based digital biomarkers to predict IR

We developed three random forest classifiers, each representing a distinct measurement modality. Feature selection was motivated by prior work demonstrating that anthropometrics, smartwatch-derived HR metrics, and CGM-derived glycemic variability indices are correlated with IR and downstream glycemic measures^9,26,27,28 29 26,27,30,31^. The first model presented an *anthropometric baseline model* comprising BMI, waist-to-hip ratio, and age (*p* = 3). These features were selected as established proxies for central adiposity and visceral fat accumulation, both independently associated with IR^43,44^. The second *smartwatch-based model* added day-night variability in heart rate and an HRV-derived daily stress score to BMI, waist-to-hip ratio, and age (*p* = 5). Reduced HRV and elevated resting HR are associated with IR through autonomic dysregulation of glucose metabolism and can be measured reliably via consumer smartwatches^26,27,28,30^. The third *CGM-based* model further incorporated CGM-derived measures, including mean and *SD* of fasting glucose, daily glucose excursions, glucose recovery time, and overall maximum glucose to BMI, waist-to-hip ratio, and age (*p* = 8). Physiologically, prior work suggests these metrics capture inter-individual differences in glycemic responses, including IR^9^ and have demonstrated predictive value for diabetes progression in non-diabetic populations ^31,32^ . For a full list of features see Table S1 and for details on model development refer to ‘Methods: Model Development’.

Briefly, model performance was evaluated in a two stage internal/external validation approach^37^, using a unified 200-iteration stratified bootstrap framework to assess uncertainty^45,37^. In Study 1 (internal validation), each iteration drew a stratified 70/30 train/test split; such that models were trained on the 70% partition (∼68 participants) and evaluated on the held-out 30% (∼29 participants). The mean estimates and corresponding 95% CIs were derived from the bootstrap performance distribution using the 2.5th and 97.5th percentiles. For Study 2 (external validation), each iteration trained a model on all (*N*=97) Study 1 participants and evaluated it on a 200-iteration stratified bootstrap resample of Study 2. Importantly, Study 2 data never entered training at any point. This unified framework ensures that internal and external confidence intervals are directly comparable across cohorts. Additionally, the 200-iteration bootstrap in the external cohort further stabilises performance estimates against sampling variability. Our primary evaluation metrics described below are derived from the Study 2 (external cohort) validation results.

### Smartwatch- and CGM-Based Digital Biomarkers Predict Insulin Resistance in an Independent Cohort

In external validation, the CGM model demonstrated the strongest discrimination, with an AU-ROC of 0.873 [0.756–0.967], AU-PRC of 0.816 [0.640–0.934] — substantially above the random-classifier base-rate baseline of 0.311 — and AU-PRG of 0.864 [0.681–0.970], reflecting 86.4% of the maximum possible precision–recall gain over a random classifier. In simple terms, an AU-ROC of 0.873 means that if we picked one individual with IR and one without at random, the model ranks the IR individual as higher risk 87.3% of the time. The smartwatch model generalized with an AU-ROC of 0.789 [0.647–0.904], AU-PRC of 0.692 [0.521–0.851], and AU-PRG of 0.690 [0.329–0.918], achieving sensitivity of 0.737 [0.526–0.933] and specificity of 0.714 [0.574–0.851] at a 0.5 decision threshold. The anthropometrics-only baseline yielded an AU-ROC of 0.749 [0.601–0.868], AU-PRC of 0.593 [0.410–0.790], and AU-PRG of 0.623 [0.300–0.860]. Full performance metrics with confidence intervals are reported in Table 1, and discrimination performance across metrics and both cohorts is illustrated in Figure 2.

**Table 1:** IR Classification Performance Metrics for Internal Development (Study 1) and External Validation on an Independent Cohort (Study 2)

| Model | Accuracy | Sensitivity | Specificity | Balanced Accuracy | Precision | F1 | AU-ROC | AU-PRC | AU-PRGC |
| --- | --- | --- | --- | --- | --- | --- | --- | --- | --- |
|  | [95% CI] | [95% CI] | [95% CI] | [95% CI] | [95% CI] | [95% CI] | [95% CI] | [95% CI] | [95% CI] |
| **Internal model development — Study 1 (N = 97)** |  |  |  |  |  |  |  |  |  |
| **CGM model (+anthropometrics)** | 0.766<br>[0.643–0.893] | 0.598<br>[0.331–0.889] | 0.845<br>[0.632–1.000] | 0.722<br>[0.567–0.865] | 0.672<br>[0.443–1.000] | 0.615<br>[0.333–0.801] | 0.822<br>[0.678–0.939] | 0.715<br>[0.470–0.894] | 0.734<br>[0.392–0.945] |
| **Smartwatch model (+anthropometrics)** | 0.745<br>[0.642–0.857] | 0.621<br>[0.333–0.889] | 0.803<br>[0.579–0.947] | 0.712<br>[0.561–0.863] | 0.615<br>[0.416–0.858] | 0.607<br>[0.399–0.800] | 0.811<br>[0.655–0.927] | 0.651<br>[0.420–0.873] | 0.693<br>[0.359–0.924] |
| **Baseline anthropometrics model** | 0.748<br>[0.607–0.893] | 0.631<br>[0.333–0.889] | 0.804<br>[0.630–0.947] | 0.717<br>[0.561–0.865] | 0.616<br>[0.416–0.858] | 0.611<br>[0.399–0.824] | 0.817<br>[0.684–0.924] | 0.648<br>[0.438–0.861] | 0.698<br>[0.399–0.921] |
| **External validation — Study 2 (N = 61, applied without retraining)** |  |  |  |  |  |  |  |  |  |
| **CGM model (+anthropometrics)** | 0.803<br>[0.705–0.902] | 0.842<br>[0.643–1.000] | 0.786<br>[0.651–0.905] | 0.814<br>[0.699–0.914] | 0.696<br>[0.500–0.875] | 0.727<br>[0.538–0.885] | 0.873<br>[0.756–0.967] | 0.816<br>[0.640–0.934] | 0.864<br>[0.681–0.970] |
| **Smartwatch model (+anthropometrics)** | 0.721<br>[0.607–0.836] | 0.737<br>[0.526–0.933] | 0.714<br>[0.574–0.851] | 0.726<br>[0.606–0.851] | 0.583<br>[0.385–0.769] | 0.622<br>[0.429–0.800] | 0.789<br>[0.647–0.904] | 0.692<br>[0.521–0.851] | 0.690<br>[0.329–0.918] |
| **Baseline model** | 0.721<br>[0.607–0.836] | 0.737<br>[0.529–0.929] | 0.714<br>[0.574–0.848] | 0.726<br>[0.602–0.845] | 0.583<br>[0.375–0.769] | 0.622<br>[0.429–0.800] | 0.749<br>[0.601–0.868] | 0.593<br>[0.410–0.790] | 0.623<br>[0.300–0.860] |

*Note.* For a list of features in each model refer to Table S1. Values are reported as mean [2.5–97.5% CI] across 200 stratified bootstrap iterations. Internal CIs: percentile interval across 200 stratified 70/30 splits of Study 1. External CIs: percentile interval across 200 bootstrap resamples of Study 2 (stratified with replacement; Study 2 data never entered training). The identical 200 seeds are applied at both stages (Study 1 & Study 2), making CIs comparable across cohorts. AU-ROC = area under the receiver operating characteristic curve (random baseline 0.5). AU-PRC = area under the precision–recall curve; random-classifier baseline equals IR prevalence (0.33 internal; 0.31 external). AU-PRG = area under the precision–recall gain curve; random-classifier baseline = 0. Balanced accuracy = (Sensitivity + Specificity) / 2. Precision = positive predictive value.

### Feature Importance

To identify the features driving IR classification in each model, we applied (SHAP) SHapley Additive exPlanations analysis to both Study 1 and Study 2 — a framework that quantifies each feature’s marginal contribution to individual model predictions, enabling both global importance ranking and directional interpretation^43,46,37,46^. See Figure S5 for feature rank stability across 200 bootstrap iterations in both cohorts. In the smartwatch model, device-derived signals — HR day-night variability and HRV-derived stress indices — accounted for approximately 18% of attributable importance (Study 1: 18.0%; Study 2: 16.7%), with BMI, WHR, and age explaining the remainder. In the CGM model, glycemic features collectively contributed approximately 46% of predictive importance (Study 1: 48.1%; Study 2: 43.4%) — including *SD* of fasting glucose, glucose recovery time, mean fasting glucose, overall maximum glucose, and excursion frequency — indicating that the model captures meaningful metabolic signal beyond anthropometrics and age alone. Feature importance rankings were highly consistent across Study 1 and Study 2 (Figure 1), supporting cross-cohort generalizability. The direction of all feature effects was consistent with established IR pathophysiology: higher BMI and WHR, greater glycemic variability, impaired post-absorptive glucose clearance, and elevated autonomic stress indices each increased predicted IR probability^9,26,27,29,30,31,32^.

**Figure 1.**
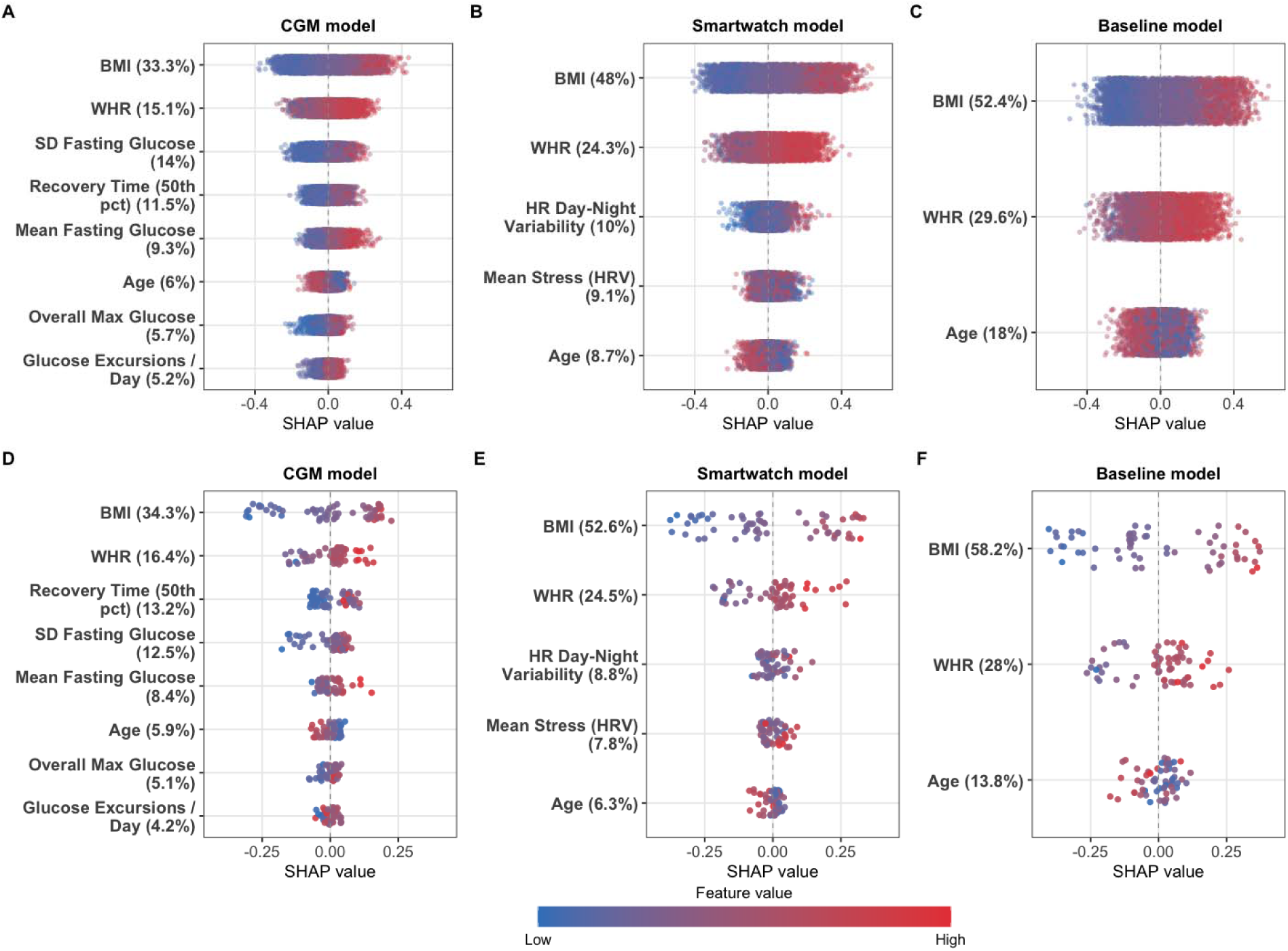
SHAP Value Distributions for the Baseline, Smartwatch, and CGM Models Across Internal and External Validation Cohorts. *Note.* The top row (A–C) shows SHAP value distributions for the internal validation cohort (Study 1; *N* = 97), where each point represents one observation from one bootstrap holdout partition, aggregated across 200 stratified 70/30 splits. The bottom row (D–F) shows corresponding distributions for the external validation cohort (Study 2; *N* = 61), where each point represents one participant with SHAP values averaged element-wise across 200 independently fitted models — each trained on all Study 1 participants using the same seeds as performance estimation, with Study 2 data never entering training at any point. Panels within each row are ordered by descending mean |SHAP| importance within each modality. The horizontal position of each point reflects its SHAP value: positive values increase the predicted probability of insulin resistance; negative values decrease it. Point colour indicates the standardised feature value (blue = low; red = high). Feature labels report each variable’s percentage share of total mean absolute SHAP importance within that model. SHAP = SHapley Additive exPlanations; CGM = continuous glucose monitor; BMI = body mass index; WHR = waist-to-hip ratio; SD = standard deviation; HR = heart rate; HRV = heart rate variability.

**Figure 2.**
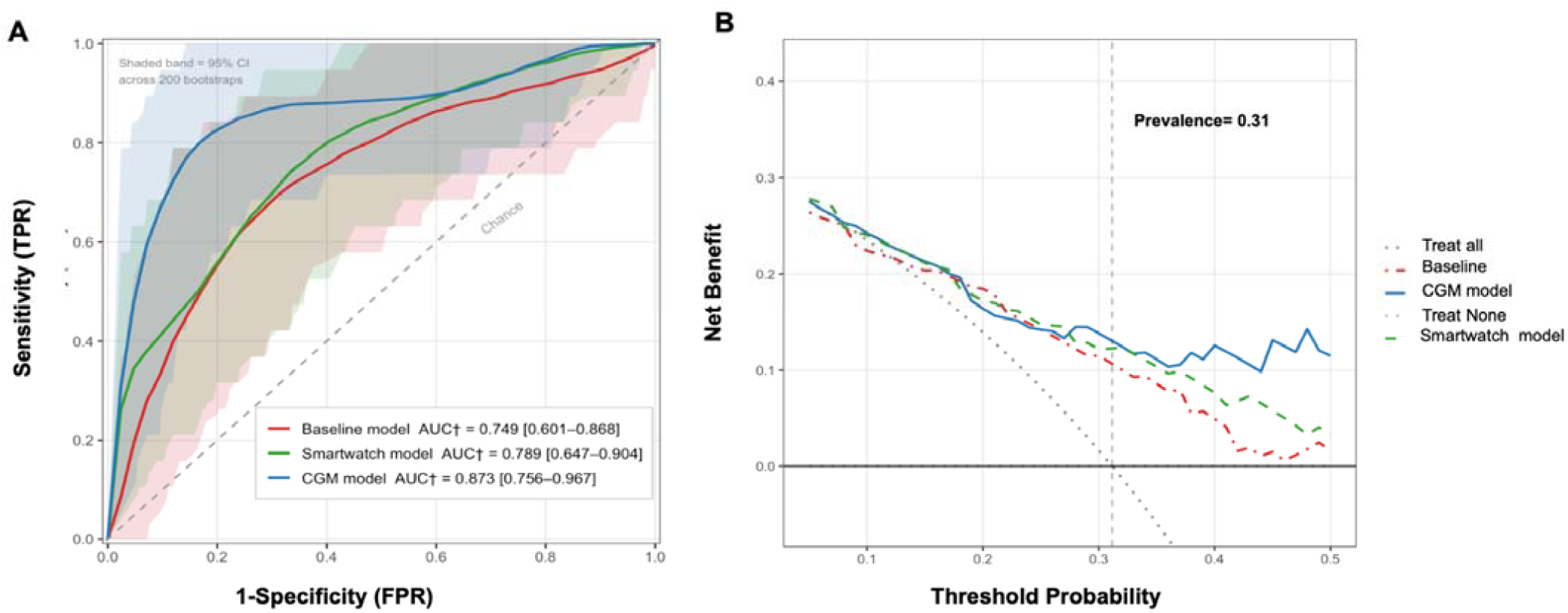
Receiver Operating Characteristic Curves and Decision Curve Analysis for the External Validation Cohort (Study 2). *Note.* Panel A displays receiver operating characteristic (ROC) curves for all three models evaluated on the external validation cohort (Study 2; *N* = 61). Each model was independently trained on all Study 1 participants (*N* = 97) across 200 iterations using independent random seeds; for each iteration, the fitted model was applied to a stratified bootstrap resample of the 61 Study 2 participants (sampled with replacement, preserving IR prevalence). Solid curves represent the pointwise mean sensitivity across the 200 per-iteration ROC curves, smoothed by isotonic regression to enforce monotonicity. Shaded bands indicate the pointwise 2.5th–97.5th percentile interval of sensitivity across the same 200 per-iteration ROC curves, capturing variability attributable to both random forest stochasticity and test-set sampling uncertainty. AUC† is reported as the mean [2.5th–97.5th percentile CI] of per-iteration AUCs, each computed on the bootstrap-resampled Study 2 subset; with values identical to those reported in Table 1. Panel B displays decision curve analysis for the same cohort, using ensemble-averaged predicted probabilities (mean across all 200 seed-specific models applied to the full Study 2 set without resampling). Net benefit is defined as TP/*N* − FP/*N* × *t*/(1−*t*), where *t* denotes the threshold probability, TP the number of true positives, FP the number of false positives, and *N* the total sample size. The dashed vertical line indicates the observed IR prevalence (*t* = 0.31). All models are compared against treat-all and treat-none reference strategies. † Monotone smoothing applied to averaged ROC curves only; AUC values are unaffected. FPR = false positive rate; TPR = true positive rate; IR = insulin resistance; CGM = continuous glucose monitor.

### Wearable-Based Models Provide Clinical Benefit Beyond Anthropometric Screening

Given that anthropometric features, particularly BMI and WHR, were the dominant predictors of IR (Figure 1), we conducted follow-up decision curve analysis (DCA) to evaluate whether the wearable-augmented models provide clinically meaningful prediction benefit beyond anthropometric features and age alone (Figure 3). DCA quantifies the trade-off between true positives and false positives across a range of clinical decision thresholds, i.e., defined as the probability at which a clinician would act on a positive prediction. Net benefit is calculated as the true-positive rate minus the false-positive rate multiplied by the odds of the threshold probability—that is, pt/(1−pt)pt/(1−pt), which reflects the relative harm of a false positive at that given decision threshold and is plotted against threshold probability. A model is clinically useful if its net benefit curve lies above both the “treat all” and “treat none” reference strategies^47^.

All three models demonstrated positive net benefit across the clinically plausible threshold range (0.10–0.45), consistently outperforming indiscriminate screening. Below the IR prevalence threshold (t = 0.31), curves were closely aligned, reflecting the dominant contribution of shared anthropometric features. At and above the observed IR prevalence, the CGM model achieved the highest net benefit (NB = 0.146), followed by the smartwatch model (NB = 0.122) and the anthropometrics-only baseline (NB = 0.115), while indiscriminate screening yielded near-zero net benefit (NB = 0.002). At the prevalence threshold, the CGM model would identify approximately 15 additional true IR cases per 100 individuals screened relative to no screening, and approximately three additional cases relative to the anthropometrics-only baseline at equivalent false-positive cost. The separation between wearable and baseline models becomes most pronounced at higher thresholds, where CGM-derived features sustain net benefit most clearly; the smartwatch model also remained above the anthropometrics-only baseline across this range (NB = 0.122 vs. 0.115 at t = 0.31, difference = 0.007), albeit with a narrower margin, while both the baseline and smartwatch curves declined more steeply beyond t = 0.35, suggesting that CGM signals may be particularly valuable in more selective, high-risk, screening contexts. Collectively, these findings suggest that free-living wearable signals carry independent metabolic information beyond body composition alone. CGM-derived features demonstrated the clearest incremental clinical utility, while smartwatch-derived (HR/HRV) signals provided a consistent, if modest, additive benefit to anthropometric IR risk stratification.

### Sensitivity Analyses

We conducted a series of follow-up analyses to assess the stability of our primary results across varying modelling specifications and analytical choices. First, to assess robustness to threshold choice, we evaluated all three models (baseline, smartwatch-based and CGM-based) at two alternative HOMA-IR cutoffs: ≥ 2.7, corresponding to the threshold proposed by Ref^48^, widely adopted in European population studies, representing borderline-to-moderate IR; and ≥ 3.5, corresponding to the 75th percentile of the internal cohort’s HOMA-IR distribution, representing more advanced IR in our data. These thresholds bracket our primary HOMA-IR cutoff of ≥ 2.9 on either side of the clinical severity continuum. Our model rank order CGM > smartwatch > baseline was preserved at both alternative thresholds in the external validation sample, closely replicating the primary findings (See Table S5 for threshold-based sensitivity analyses). Next, we assessed feature rank stability across 200 bootstrap runs (Figure S5); and compared external validation model performance across different imputation methods (Table S6). Across both robustness checks, the results closely parallel those presented in the main manuscript.

## Discussion

Insulin resistance affects approximately one in four adults globally and is the primary pathophysiological driver of type-2 diabetes, cardiovascular disease, and metabolic syndrome^49–51^. Critically, IR can precede overt type-2 diabetes by up to a decade to 15 years, yet is typically identified reactively—via venepuncture-based blood tests, and often through incidental screening. Relatedly, many individuals remain unaware until downstream complications prompt clinical investigation. Digital biomarkers, defined as continuously sampled physiological signals derived from wearables^52^, present new opportunities to shift IR screening from reactive, clinic-based testing toward proactive, remote screening as individuals go about their daily lives. Unlike clinical lab tests, continuous monitoring can enable dynamic risk stratification by capturing temporal physiological fluctuations in real-world conditions, and beyond single-timepoint measurements.

To this end, we evaluated whether AI-algorithms applied to digital biomarkers derived from consumer smartwatches—tracking heart rate and heart rate variability, and CGMs—tracking fasting glucose, excursions, and recovery time, could detect IR in free-living conditions. Both smartwatch and CGM-based digital biomarker models can feasibly predict IR in an independent cohort without retraining, relative to an anthropometrics-only baseline (AU-ROC (AU-ROC = 0.749, AU-PRC = 0.593). Specifically, the CGM-based model achieved an AU-ROC of 0.873 [0.756–0.967] and AU-PRC of 0.816 [0.640–0.934], and the smartwatch-based model achieved an AU-ROC of 0.789 [0.647–0.904] and AU-PRC of 0.692 [0.521–0.851]. These results provide proof-of-concept evidence that passively collected, theory-driven wearable signals can capture physiologically meaningful signals for IR detection; critically among adults with normoglycemia and prediabetes, prior to formal type-2 diabetes diagnosis. Our findings support the feasibility of early, wearable-based population screening without venepuncture or laboratory processing.

Importantly, our findings represent several key advancements when contextualized against existing work. To our knowledge, no studies have used free-living CGM data without structured glucose tests for formal IR prediction. Only one recent study has employed consumer smartwatches for IR prediction in free-living conditions, the seminal WEAR-ME study published in *Nature* in 2026, which validated its models in an independent sample of 72 participants, comparable to our external validation sample (Study 2; *N* = 61). Critically, to achieve a blood-test-free AU-ROC of 0.75, the WEAR-ME study required a wearable foundation model pretrained on 40 million hours of sensor data. Reaching an AUC of 0.88 additionally required fasting glucose and a full lipid panel^33^, constraining scalability and repeated, population-level reach. A prior CGM-based study likewise reached an AU-ROC of 0.88, through a structured at-home oral glucose tolerance test^31^. Critically, our models match or exceed these benchmarks under substantially more accessible conditions — no lab tests, no structured protocols, and no large-scale pretraining — using passively collected, free-living data and a small, theory-driven interpretable feature set. That comparable performance is achievable with a parsimonious, theory-driven feature set suggests that complex multimodal pipelines may not be necessary for initial IR screening, meaningfully lowering the practical burden of data collection and model deployment in real-world settings.

Relatedly, using theory-driven, interpretable features addresses a critical gap in building clinician trust^34^. Post-hoc SHAP explanations revealed that our wearable-based predictors strongly align with known IR pathophysiology. For the CGM model, elevated mean fasting glucose and delayed postprandial glucose recovery were key predictors, reflecting impaired hepatic overnight glucose suppression and slowed peripheral tissue uptake^9^. However, our feature results also provide actionable design implications for over-the-counter CGM device platforms. The glucose features that best predicted insulin resistance in our data, e.g., day-to-day fasting glucose variability and post-meal glucose recovery, are critically absent from summary dashboards of existing CGM apps, which tend to display summaries such as mean glucose and time in range, that are traditionally designed for diabetes management. Importantly, the insulin resistance predictive features we identified require no new hardware: they can be computed from the same raw signal already collected by over-the-counter CGM devices. Integrating these interpretable features into over-the-counter CGM platforms in future work, can extend their utility from diabetes management to preventative- insulin resistance screening, for individuals without formal type-2 diabetes diagnoses. This represents a key opportunity for a paradigm shift in how CGM data can be reported and interpreted for non-diabetic populations: rather than repurposing metrics designed to manage overt hyperglycemia, CGM platforms can fruitfully develop and test a new layer of physiology informed metrics, purposefully engineered to detect early metabolic dysfunction in daily life.

In the smartwatch model, elevated daily stress and day-to-night variability of day-night heart rate were similarly important drivers. These features capture the autonomic dysregulation typical of IR, specifically the persistent sympathetic overactivation that blunts restorative nocturnal parasympathetic dominance^29,30^. Notably, follow-up analyses showed that combining CGM and smartwatch (HR/HRV) features did not meaningfully improve IR predictability (Table S7). This finding may partly reflect that adding more features increases dimensionality without a proportional increase in observations, making it harder to detect incremental gains in discriminative performance. It may also reflect plausible physiological overlap between the two signal streams: both glucose dysregulation and autonomic dysfunction are downstream consequences of IR, such that once CGM captures the glycemic signal, HR/HRV may carry limited additional discriminative information. However, replication in larger cohorts is needed to disentangle these plausible explanations.

Interestingly, adding common physical-activity features, such as mean step count or activity minutes, did not independently improve model performance beyond the HR/HRV-based features, suggesting that the autonomic features sampled here may capture deeper intrinsic physiological relationships (Supplement B: Physical Activity Analyses and Table S7). Another possibility is that the 10-day monitoring window may be too short to obtain stable and representative habitual activity patterns. Consistently, we observed low agreement between self-reported and smartwatch-measured activity in both cohorts (maximum *r* = −0.37 between self-reported activity and either mean total steps or mean active minutes across cohorts). This observation may also explain the divergent findings with the WEAR-ME study, where physical activity features meaningfully contributed to IR prediction in the context of considerably longer monitoring duration [up to 90 days vs. approximately 10 days in the present study]. Interestingly, the WEAR-ME study also lacked waist-to-hip measurements, a known predictor of IR^43,44^. which may absorb a portion of the variance that physical activity features may otherwise capture^53^. Overall, whether a longer activity monitoring window would restore an independent contribution from physical activity features warrants future investigation.

One defining strength of this study is the use of an external independent validation cohort and the models’ generalization to a geographically distinct cohort without any retraining. This stability held despite notable baseline differences between the internal and external cohorts regarding fasting glucose, BMI, directional trends in prediabetes prevalence, and device wear days (See Supplement A and Table S2 for cohort descriptives). This robustness suggests the wearable-based algorithms developed here can successfully isolate underlying metabolic signals rather than overfitting to site-specific characteristics. In other words, future population deployments would not require constant, costly local recalibrations. A further strength is that our DCA analyses showed that wearable-derived features carry independent predictive benefit beyond conventional anthropometric factors alone (Figure 3). At the observed IR prevalence threshold (*t* = 0.31), the CGM model achieved the highest net benefit, corresponding to approximately three additional true IR cases identified per 100 individuals screened relative to the anthropometrics-only baseline at equivalent false-positive cost. The smartwatch model provided a consistent, if more modest, additive benefit. The separation between wearable and baseline models was most pronounced at higher thresholds, where CGM-derived features sustained net benefit most clearly, suggesting more value in higher-risk or more selective screening contexts. Together, these findings build our confidence that the addition of remote, low-burden, wearable features can meaningfully augment anthropometric IR risk stratification.

Clinically, the digital biomarker models developed here provide a promising early stage infrastructure for scalable, population-level IR screening. Considering the estimated 560 million global smartwatch users^22^ and the expanding over-the-counter CGM market^24^, a 10-day passive monitoring period could automatically flag elevated IR probability and prompt clinical follow-up, effectively reducing the *time-to-diagnosis* gap. The clinical value of early detection is substantial: IR often precedes a type-2 diabetes diagnosis by 10 to 15 years^12^. Identifying metabolic dysfunction during this early window enables targeted lifestyle interventions, which are 44% more likely to reverse prediabetes to normoglycemia compared to usual care^14,15^. Another strength of the models presented here is flexibility suited to available device data. While CGM-based models performed best in our data (AU-ROC=0.87), smartwatch-based screening may (AU-ROC=0.79) provide a less invasive option that may improve patient acceptance and screening uptake, particularly in populations where CGMs may be initially unavailable. Device interchangeability may allow model deployment in diverse settings, utilizing the device data that a particular patient has available and thus broadening the potential screening reach.

In practice, such an IR screening system could operate as a continuously updating background process on a consumer smartwatch or CGM device. After an individual initially provides basic anthropometric data, the system could periodically recompute IR risk from passively accumulated HR, HRV, or CGM features without requiring further user input or on-site visits. Elevated IR risk estimates could then trigger a soft alert prompting the user to seek confirmatory insulin and glucose blood tests, effectively functioning as a first-line triage layer between population-level screening and confirmatory clinical diagnosis. Importantly, we do not position CGM-based IR screening as a lower-cost replacement for HOMA-IR. Rather, its value lies in passive, longitudinal risk detection from data that can be increasingly generated outside clinical care as over-the-counter CGMs become broadly accessible. Remote IR screens could direct at-risk-individuals to confirmatory fasting glucose and insulin testing, who otherwise would not get tested. The premise of our digital biomarkers is to substantially increase uptake of confirmatory HOMA-IR testing by identifying at-risk individuals, earlier and at scale. When deploying these models in practice, it is important to tailor the classification threshold—the predicted probability above which an individual is flagged as having IR. In our development cohort, a standard 0.5 probability threshold yields balanced sensitivity and specificity. However, in low-prevalence populations (e.g., normoglycemic adults), lowering the threshold (e.g., to 0.3) may improve sensitivity at the cost of more false positives, ensuring fewer missed cases. Conversely, in high-prevalence settings (e.g., prediabetic cohorts), raising the threshold (e.g., to 0.6) would increase positive predictive value, reducing unnecessary confirmatory testing. Threshold selection may prioritize the metric most aligned with context-specific clinician goals: sensitivity for catch-all screening, specificity for ruling out low-risk individuals, or positive predictive value when confirmatory HOMA-IR testing is resource-limited.

Beyond model threshold calibration, IR risk may also be continuously adapted in practice. We applied a HOMA-IR ≥ 2.9 threshold based on comparability with prior work^33,54^, but optimal cutoffs vary by ethnicity, sex, and age^54^. For example, Asian populations may show IR-related metabolic dysfunction at lower HOMA-IR values than European populations, with some studies suggesting ethnicity-specific thresholds (e.g., ≥2.0 for Asian vs ≥2.9 for European cohorts) to improve clinical relevance^54,55^. Similarly, HOMA-IR naturally increases with age, such that a fixed threshold may over-diagnose IR in older adults while missing cases in younger individuals. Our models could thus be recalibrated using age-stratified, sex, or ethnicity-specific HOMA-IR cutoffs. Integrated within existing public health infrastructure, such as primary care electronic health records, this approach could enable scalable, equitable early detection of IR in settings where laboratory access is limited or where at-risk individuals would not otherwise present for testing. However, it is also important to note that digital screening may inherently risk widening health disparities if deployment is not equity-conscious. While the underlying AI-READI dataset was purposefully designed to capture diverse clinical sites^35^, smartwatch and CGM device ownership remains skewed toward higher-income populations. Extending this paradigm to lower-resource settings will require targeted interventions, such as subsidized devices, and continuous algorithm validation across socioeconomically diverse cohorts to prevent bias.

In this direction, emerging health-economic evidence suggests that preventive, digital biomarker–based screening for prediabetes can be cost-effective at a population scale. For example, a recent Markov-modeling study from the Swiss payer perspective found digital biomarker-based screening for prediabetes (with 70% sensitivity and 70% specificity) to be highly cost-effective compared to current opportunistic approaches^56^, but requires reimbursement models that align payer incentives with upfront investment costs^57^. Value-based or outcome-based reimbursement— rewarding prevention success (i.e., reduction of IR), rather than discrete clinical visits, is a promising pathway to sustain continuous, real-world monitoring programs^58,59^, strengthening the translational rationale for the proposed pipeline.

Several limitations must be noted. Our sample sizes (*N* = 97 in Study 1 and *N* = 61 in Study 2) are sufficient for proof-of-concept but remain underpowered for granular subgroup analyses and for data-driven (“kitchen sink”) feature selection involving a broader, more comprehensive set of potentially clinically relevant predictors. Counterintuitively, we observed lower internal (Study 1) than external performance (Study 2) across models (Figure 1). This observation may, in part, reflect the fact that internal evaluation was conducted on the held-out 30% test set (n∼29) across 200 bootstrap splits. This internal evaluation context is likely to yield higher variance than the external cohort (*N* = 61), which was evaluated in full on each iteration. Relatedly, the closer alignment between smartwatch and baseline models observed internally (Study 1) — but not externally (Study 2) — may similarly reflect the limited statistical resolution of a ∼29-patient test set rather than a true absence of performance differences. These observed differences may also partly reflect the higher proportion (+4.2 percentage points, ∼8% relative increase) of participants with prediabetes in Study 2 vs. Study 1, which could strengthen the detectable signal (Study 1: 51.5% prediabetic, n = 50 vs. Study 2: 55.7% prediabetic, n = 34). Critically, we did not observe a model performance drop from internal to external validation, which argues against overfitting, and builds our confidence that the models generalized beyond the derivation sample. We relied on HOMA-IR as our ground-truth classification, which, while clinically established, is a surrogate estimate and is less precise than the hyperinsulinemic-euglycemic clamp^17,18^. Information on sex and ethnicity was unavailable in both studies, precluding inclusion as a predictive IR feature and stratified sub-group analyses of IR prediction. The lack of sex and ethnicity data is an important limitation given established sex differences in insulin sensitivity, visceral adiposity distribution, and hormonal modulation of glucose metabolism^60^, and varying ethnicity-based HOMA-IR cut-offs^54,55^. However, although proposed HOMA-IR cut-offs vary across populations, we observed broadly parallel results across different thresholds (Table S5). A further limitation is that the smartwatch-derived “stress” feature relies on a Garmin proprietary algorithm, which is not fully transparent. Future prospective digital biomarker studies combining consumer wearables with validated research-grade devices^61^, may fruitfully disentangle the contribution of device-specific processing from underlying physiological signals, and establish whether raw and open-source feature pipelines can yield comparable predictive signals. Finally, the cross-sectional design precludes the longitudinal tracking of IR changes. Prospective designs in which continuous wearable monitoring precedes clinical HOMA-IR assessment will be essential to determine the degree to which digital biomarkers can offer an early-warning advantage.

Future research is needed to evaluate these digital biomarkers in larger and more diverse cohorts, particularly non-Western populations, and those across varying socioeconomic profiles, to confirm their global applicability. Further work should also assess model generalizability across different IR risk profiles, including varying proportions of normoglycaemia and prediabetes. We observed a relatively balanced distribution (Study 1: 51.5% prediabetic (HbA1c > 5.7), *n* = 50; Study 2: 55.7%, *n* = 34). However, our sample sizes do not permit evaluation of model performance under different population prevalences. Further work is therefore needed to assess model stability and generalizability across cohorts with differing baseline risk distributions. Longitudinal validation is also a critical next step to determine whether these models can accurately track dynamic, within-person changes in IR in response to structured lifestyle interventions. Future computational efforts should explore a direct head-to-head comparison with large foundation models on identical datasets, alongside the integration of these predictive frameworks into LLM-based recommendation systems for personalized behavioral coaching. Finally, comprehensive health economic analyses of this digital screening pipeline are needed across diverse healthcare and resource settings, including both low- and high-income contexts. Scaled to approximately 560M smartwatch users and an emerging market of non-diabetic CGM users, our decision-curve results (∼3 additional true IR cases per 100 screened) potentially correspond to ∼9–13M additional early IR detections, assuming a 15–30% prevalence among smartwatch and CGM users. However, prospective validation in large real-world cohorts remains essential to confirm these projections and to better inform screening implementation and reimbursement strategies across heterogeneous health systems.

In conclusion, this study is among the first to demonstrate that passively collected, free-living wearable signals — derived from CGM and consumer smartwatch devices — can feasibly detect IR in individuals with normoglycemia and prediabetes, generalizing to an independent, geographically distinct cohort without retraining. Critically, our models operate precisely within a critical clinical window: the decade or more preceding overt glucose dysregulation during which IR is present but conventional screening would not yet trigger intervention. Using small, interpretable feature sets grounded in established IR pathophysiology, we establish an early foundation for scalable, population-level IR screening that can operate continuously in the background of daily life — requiring no clinical visit, no blood draw, and no active user burden. As wearable adoption continues to expand globally, integrating AI-driven IR risk assessment into consumer health platforms represents a tractable and timely pathway toward earlier identification of at-risk individuals, with potential to substantially reduce population-level burden of type-2 diabetes and downstream complications.

## Methods

### Data

We used publicly available data from the AI-READI project^36,41,42^, an initiative funded by the National Institutes of Health under the Bridge2AI Program. The AI-READI dataset was designed to support machine learning research on type-2 diabetes, with a focus on predicting disease development and progression. We used the second release of the dataset (v2.0.0), made publicly available in November 2024.

### Ethics

The original data collection was approved by the University of Washington Institutional Review Board (IRB) on December 20, 2022, and all participants provided informed consent in accordance with relevant ethical guidelines (IRB Study Number: 00016228; FWA Number: #00006878). The present secondary analysis of these publicly available data was reviewed by the Ethics Committee of the University of St. Gallen (HSG) in St. Gallen, Switzerland, which granted an exemption confirming that the study meets applicable ethical standards. The current investigation only made use of de-identified data, maintaining anonymity of the original participants.

### Participant recruitment

As part of the original investigation, participants were recruited across three sites in the United States: Birmingham, Alabama (University of Alabama at Birmingham); San Diego, California (University of California San Diego); and Seattle, Washington (University of Washington). For full details on recruitment procedures and eligibility criteria, refer to Refs. ^36,41,42^. Eligible participants were adults aged 40 or older who could provide informed consent and communicate in English. Individuals who were pregnant or had gestational or type 1 diabetes were deemed ineligible. Recruitment pools were identified by screening Electronic Health Records for diabetes- and prediabetes-related ICD-10 codes among individuals who had encountered the sites’ health systems within the prior two years. Data collection for the second version of the AI-READI dataset, used in the present study, spanned July 19, 2023 to July 31, 2024.

### Study procedure

The AI-READI study protocol comprised three phases: pre-visit questionnaires, an on-site data collection visit, and a post-visit remote data collection period. Prior to the clinic visit, participants completed questionnaires capturing demographic and background information, including age. During the on-site visit, participants underwent anthropometric measurements — including height, weight, and waist and hip circumference — and provided blood samples for fasting glucose, fasting insulin, HbA1c; with fasting glucose and fasting insulin constituting the primary clinical variables used to calculate HOMA-IR in the present analyses. Additional assessments included retinal imaging, electrocardiography, and cognitive function evaluation, beyond the scope of the present study. Following the clinic visit, participants entered an approximate 10-day wearable data collection phase, during which they wore a Dexcom G6 CGM and a Garmin Vivosmart 5 fitness tracker. Participants received $200 compensation for their time and involvement.

### Participants

The starting dataset comprised 1,067 participants from the AI-READI dataset. The full step-by-step participant flow, following PRISMA guidelines, is reported in Figure S1. Participants dependent on insulin therapy were first excluded (*n* = 130), as exogenous insulin administration directly confounds HOMA-IR estimation and may alter glycemic dynamics captured by CGM in ways that reflect treatment effects rather than underlying metabolic physiology and lifestyle factors. Participants who self-reported not fasting for at least 8 hours prior to blood collection were further excluded (*n* = 680), as the glucose and insulin values required for HOMA-IR computation^18^ are contingent on a fasted state. Participants with a type-2 diagnosis (based on HbA1c ≥ 6.5%) were subsequently excluded (n = 62), leaving 195 participants below the diagnostic threshold. This restriction was intentional as we specifically aim to develop digital biomarkers for insulin resistance screening from a prevention-oriented perspective, targeting individuals who have not yet received a formal type-2 diabetes diagnosis and whose metabolic profiles are less likely to be confounded by pharmacological treatment, clinical management decisions, or behavioral modifications prompted by a known diagnosis.

Two parallel analytical pipelines were pursued, as illustrated in Figure S1. For Study 1 (Model Development and Testing), participants from the University of Washington (Seattle, WA) and the University of California San Diego (San Diego, CA) were retained for model development (n = 123). The site at the University of Alabama at Birmingham (UAB; Birmingham, AL) was reserved as an independent holdout for external validation (Study 2). Participants lacking wearable (CGM and/or smartwatch) data (n = 24), and who provided implausible HOMA-IR values, i.e., indicative of a failure to observe the required minimum 8-hour fast (*n* =2) were excluded (See Figure S4 for HOMA-IR outlier exclusion and the corresponding distributions before and after removal), yielding a final analytic sample of 97 participants for Study 1. For Study 2 (External Validation), only participants from the UAB site were retained (*n* = 72), ensuring a non-overlapping, geographically distinct validation sample. After excluding participants with missing wearable (CGM and/or smartwatch) data (*n* = 9), unavailable blood samples (n = 1), and implausible HOMA-IR values (*n =1*), the final analytic sample for Study 2 included 61 participants.

### Sample size considerations

Given that the present study used available data from the parent AI-READI project, no formal a priori power calculation was conducted. The analytic sample was determined entirely by data availability following the application of eligibility and quality criteria. The resulting sample sizes (Study 1: *N* = 97; Study 2: *N* = 61) are comparable, or relatively larger, than those reported in prior studies employing wearable-derived features for glycemic dysregulation prediction^31,62^ and multimodal prediction in digital health applications^37^. To build confidence in the stability and generalizability of findings despite the limited sample, several steps were taken: (1) external validation in an independent, geographically distinct cohort (Study 2) to assess generalizability and mitigate overfitting; (2) bootstrap resampling to estimate uncertainty in performance metrics; and (3) full reporting of estimates with confidence intervals to support interpretation.

### Measures

We used three feature modalities for model development: (1) anthropometric and demographic variables derived from clinical assessments and pre-visit questionnaires, (2) glycemic features derived from CGM recordings, and (3) cardiovascular and autonomic features derived from a wrist-worn consumer smartwatch (Table S1). Additional analyses with smartwatch-derived physical features are reported in Supplement B.

#### Baseline body composition and demographic measures

Three anthropometric and demographic variables were derived from the on-site clinical visit and pre-visit questionnaire: body mass index (BMI; kg/m²) was calculated from measured height and weight during the in-person visit. Waist-to-hip ratio (WHR) was computed by dividing waist circumference by hip circumference, both measured in centimeters, providing an index of central adiposity. Participant age (years) was obtained from the pre-visit questionnaire.

#### Smartwatch-based (HR/HRV) measures

Two cardiovascular and autonomic features were derived from the Garmin Vivosmart 5 wrist-worn device over the free-living monitoring period. HR Day–Night Variability was operationalized as the standard deviation of the difference between daytime and nocturnal heart rate across the entire monitoring period, reflecting the day-to-day consistency of circadian autonomic regulation. Greater variability indicates less stable nocturnal parasympathetic dominance, which has been associated with sympathetic overactivation and cardiometabolic dysregulation^63^. The HRV-based ‘Daily Stress’ score presents a continuous composite score (range 0–100), which quantifying beat-to-beat intervals in heart rate variability as a proxy for sympathetic–parasympathetic balance of the autonomic nervous system — with lower HRV indicating sympathetic dominance and higher perceived stress load^64^. The HRV-based estimate of physiological stress was averaged daily across the entire monitoring period.

#### CGM-based measures

Five glycemic features were derived over the approximately 10-day free-living monitoring phase using the Dexcom G6, selected to capture distinct theory-driven dimensions of glycemic physiology: habitual fasting glucose level, day-to-day fasting instability, excursion frequency, speed of glucose clearance, and acute peak hyperglycaemia^32,65^. For feature correlations, see Table S4; no feature correlation surpassed (*r* = −0.57) suggesting no high multicollinearity exists in the full feature set. Mean Fasting Glucose was computed as the mean of daily mean interstitial glucose values within the wake-to-first-meal window, where meal onset was detected as the first time point at which glucose rose ≥15 mg/dL above the wake baseline across two consecutive readings within 15 minutes^66^. *SD* Fasting Glucose was computed as the standard deviation of these daily mean fasting glucose values across monitoring days, indexing day-to-day instability in the pre-rise glycemic state, consistent with Ref^66^. Excursions per day was computed as the mean number of daytime glycemic excursions per monitoring day, where an excursion was defined as a glucose rise ≥15 mg/dL above a rolling local baseline^38,67^, confirmed by two consecutive readings within 15 minutes, computed over daytime hours (06:00–22:00). Fifty % Recovery Time refers to the median time (minutes) from excursion peak to 50% recovery of the glucose rise across all daytime excursions, serving as a proxy for the efficiency of glucose clearance^68^. Overall Peak Glucose captures the maximum interstitial glucose value (i.e., excursion) recorded throughout the monitoring period, indicating a person-specific upper bound of glycemic excursions^69^.

### Ground truth IR classification

IR status was determined from fasting plasma glucose and insulin samples collected at the in-person study visit. A continuous insulin resistance score was first computed using the HOMA-IR equation^18^ (HOMA-IR formula presented in Figure S3). Values indicative of a failure to observe the required minimum 8-hour fast were excluded (HOMA-IR > 35; median + *3SD*), resulting in the removal of (n=2) participants in Study 1 and (n=1) in Study 2. See Figure S4 for the corresponding HOMA-IR distributions before and after outlier exclusion. Consistent with Refs. ^33,54^, IR was binarised using a HOMA-IR cut-off of 2.9, classifying participants scoring at or above 2.9 as having IR (class 1); and those below as Non-IR (class 0). This threshold was selected for comparability with Ref.^.33,^ derived from a US population with a closely matching demographic and metabolic profile (median BMI = 28 kg/m², median age = 45 years, median HbA1c = 5.4%), closely resembling the present cohorts (Study 1: median age = 57 years, median BMI = 28.1 kg/m², median HbA1c = 5.7%; Study 2: median age = 56 years, median BMI = 31.2 kg/m², median HbA1c = 5.7%). Applying the 2.9 threshold yielded 32 IR and 65 Non-IR participants in Study 1 (IR prevalence = 33%) and 19 IR and 42 Non-IR in Study 2 (IR prevalence = 31%). To assess robustness to threshold choice, model performance was additionally evaluated across alternative HOMA-IR cut-offs (Table S5).

### Data Preparation

#### Smartwatch Data Pre-processing

Prior to feature extraction, raw heart rate data from the Garmin Vivosmart 5 were cleaned as follows. Readings below 30 bpm or above 220 bpm were removed as artefact. Features derived as HR aggregates were computed for any time window containing at least one valid reading (e.g, zero values were treated as missing, as these reflect sensor dropout rather than a physiological reading). Day–night HR difference summaries required a valid sleep window derived from the sleep onset and offset derived from the Garmin sleep stage recordings. Where no sleep window was identified for a given day, that day was excluded from the day–night variability calculation. HR Day–Night Variability was then computed as the standard deviation of the daily differences between mean daytime HR and mean nocturnal HR (HR_day − HR_night) across the monitoring period. A minimum of two valid daily differences was required for the *SD* to be computable. The absence of a valid Garmin-detected sleep window accounted for all HR day–night variability missingness, affecting *n*=7 of *n*=97 participants (7.2%) in Study 1 and *n*=2 of 61 participants (3.3%) in Study 2. Mean Daily Stress was computed by averaging the daily Garmin-derived HRV stress score across all valid monitoring days, with (‘-1’, and ‘-2’) values indicating no signal removed. Mean Daily Stress was unavailable for n=1 of 97 Study 1 participants (1%), attributable to insufficient valid Garmin monitoring days. No Participants had missing Mean Daily Stress values in Study 2.

#### CGM Data Pre-processing

Prior to feature extraction, raw CGM data were cleaned by removing readings outside the Dexcom G6 operating range (40–400 mg/dL) and excluding monitoring days with fewer than 230 of 288 expected five-minute readings (80% coverage threshold). Timezone-aware local time was assigned to all readings using participant-specific timezone data derived from Garmin sleep tracking, enabling accurate daytime windowing and fasting window computation. The resulting cleaned time series formed the basis for all five CGM-derived features. Feature-level missingness was low and arose from three causes: 6 of 97 Study 1 participants (6.2%) and 2 of 61 Study 2 participants (3.3%) lacked valid wake timestamps, preventing fasting window definition entirely and producing missing values for both fasting glucose features; an additional 4 of 97 Study 1 (4.1%) and 2 of 61 Study 2 (3.3%) participants had only a single valid fasting day, yielding a computable mean but an undefined *SD*; and 3 of 97 Study 1 participants (3.1%) had insufficient daytime CGM coverage across all monitoring days, precluding excursion and recovery time computation. In total, missing values were present for mean fasting glucose in n=6 of n=97 (6.2%) Study 1 and n=2 of n=61 (3.3%) Study 2 participants. *SD* fasting glucose values were unavailable for n=10 of n=97 (10.3%) participants in Study 1 and *n=4* of n=61 (6.6%) in Study 2. Excursions per day and 50% recovery time each, were missing for n=3 of *n=97* (3.1%) participants in Study 1 and Overall peak glucose for *n=1* of 97 (1.0%). No missing values were observed for these variables in Study 2.

Following feature extraction and preprocessing, two separate analysis-ready datasets were constructed for Study 1 (*N* = 97) and Study 2 (*N* = 61), serving as the internal development and external validation sets, respectively. Anthropometric features (BMI, age, WHR) and mean daily stress were fully observed in both samples. Missingness specific to CGM-derived and HR-based features was low in magnitude (Study 1: range 1.0–10.3%; Study 2: range 0–6.6%), and was specifically attributable to absent wake or sleep timestamps, insufficient valid monitoring days, and inadequate sensor coverage — as described in the preprocessing sections above. All missing values were handled by median imputation, with imputation parameters estimated exclusively from the training partition (70% of Study 1) and applied without re-fitting to the internal holdout and external validation sets to prevent data leakage. Results remained highly consistent across alternative imputation strategies, particularly k-nearest neighbors (*k* = 3, 5, 7) (Table S6).

### Model Development

Drawing on recent work on multimodal prediction with similar sample sizes^37^, we selected random forest as the primary modelling algorithm. Random forests accommodate non-linear relationships and feature interactions without distributional assumptions, are robust in small samples with moderate feature sets, and yield interpretable feature importance estimates amenable to SHAP-based decomposition^70,71^ and are commonly adopted in small-sample digital health prediction studies (for example Refs. ^37,72^). We developed three separate random forest classifiers to predict IR (HOMA-IR ≥ 2.9), each incorporating a different predictor set: an anthropometric reference model (Baseline Model: BMI, age, WHR), a smartwatch-derived model (Smartwatch Model: mean daily stress, HR day–night variability, age, BMI, WHR), and a CGM-derived model (CGM Model: mean fasting glucose, SD fasting glucose, excursions per day, 50% recovery time, overall peak glucose, BMI, age, WHR). Refer to Table S1 for a full list of features used in modeling.

All models were trained on Study 1 (*N* = 97) using a stratified 70/30 train–holdout split at the individual level, ensuring that all data from a given participant appeared in only one partition, thereby preventing any individual-level leakage. Hyperparameters were fixed a priori across all models per Breiman’s default; CGM: *mtry* = 2, smartwatch: *mtry* = 2, baseline: *mtry* = 1 (300 trees; *mtry* = ⌊√p⌋; nodesize = 1)^70^, eliminating the need for hyperparameter tuning or an inner cross-validation loop. To evaluate model stability, we performed 200 bootstrap resamples of the training data. For each bootstrap sample, models were refitted using the same fixed hyperparameters. Class imbalance (IR prevalence ≈ 32% across both studies) was addressed via random upsampling of the minority class within each bootstrap training set prior to model fitting^73^. Missing predictor values were imputed using median imputation, with imputation parameters estimated exclusively from the training data within each resample to avoid information leakage. See Table S6 for results across additional imputation strategies. For each iteration, we recorded AU-ROC, AU-PRC, and AU-PRG, along with standard classification metrics (accuracy, sensitivity, specificity, precision, F1). Internal (Study 1) 95% confidence intervals were derived from the 2.5–97.5 percentiles across the 200 bootstrap iterations.

### Model Evaluation

For external validation, each of the three models was separately applied to Study 2 data (n = 61) without any retraining. To obtain stable performance estimates and confidence intervals comparable to those from internal validation on Study 1, we repeated the evaluation across 200 iterations of bootstrapped resamples. In each iteration, a new random forest was trained on the complete Study 1 dataset (*N* = 97), with the same fixed hyperparameters, median imputation, and stratified upsampling procedure applied during internal validation. The resulting model was then used to generate predicted IR probabilities for all 61 Study 2 participants, yielding a set of classification metrics (sensitivity, specificity, etc.) and AUC-ROC, AU-PRC, and AU-PRG per iteration. These metrics were subsequently aggregated as the mean and 2.5–97.5th percentiles to calculate the external 95% CIs, over 200 bootstrap iterations. Similarly, we report mean of the bootstrapped SHAP values, averaged element-wise, to ensure that reported feature importances reflect the stable central tendency across all 200 fitted models rather than any single random forest.

## Data Availability

The data analyzed in this study are derived from the Flagship Dataset of Type 2 Diabetes from the AI-READI Project (v2.0.0), publicly available on the FAIRhub data platform (https://fairhub.io/datasets/2). Full documentation of the dataset structure, data modalities, and access procedures is available at https://docs.aireadi.org/docs/2/about.

## Code Availability

Scripts to reproduce the main analyses are available at: https://github.com/miajov/IR-Prediction

## Author Contribution

Conceptualization: Author, M.M., M.B., F.W., R.S., Investigation: M.J., Data curation: M.J., V.B., R.S., Q.J., Formal analysis: M.J., M.F., V.B., M.B., R.S., Q.J. Funding: M.J. Writing—original draft: M.J., M.M. Writing—review and editing: M.J., M.M., M.F., M.B., R.S., F.W., M.M., Q.J.

## Funding Acknowledgements

M.J., M.F, V.B, Q.J., affiliated with the Centre for Digital Health Interventions (CDHI), a joint initiative of the Institute for Implementation Science in Health Care, University of Zurich, the Department of Management, Technology, and Economics at ETH Zurich, and the Institute of Technology Management and School of Medicine at the University of St.Gallen. CDHI is funded in part by MavieNext, an Austrian healthcare provider, CSS, a Swiss health insurer, and MTIP, a growth equity firm.

## Supplements

**Figure S1.**
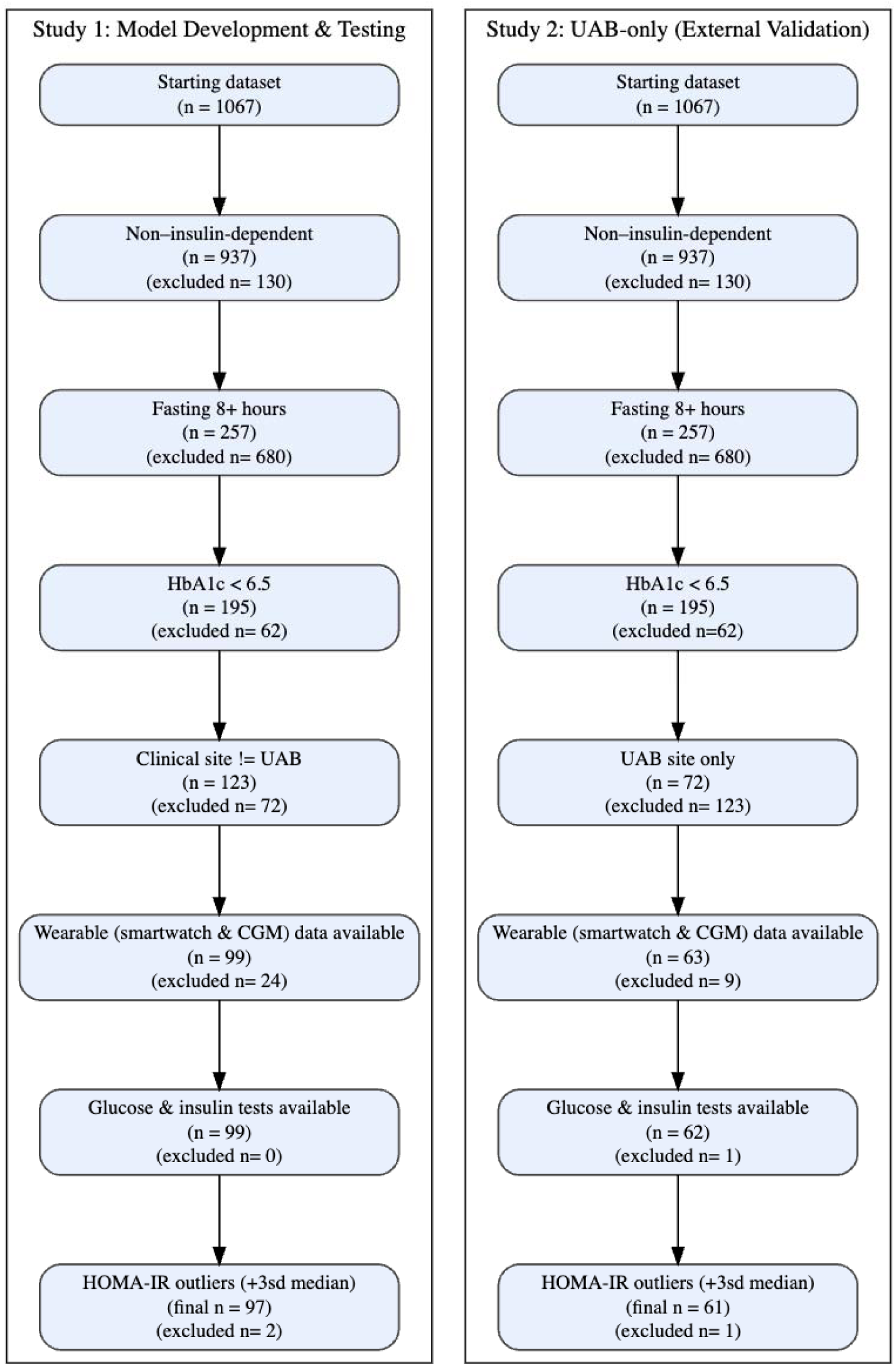
Participant flow chart across Study 1 and Study 2. *Note*. The diagram illustrates the number of participants included or excluded at each stage of data collection and pre-processing.

**Figure S2.**
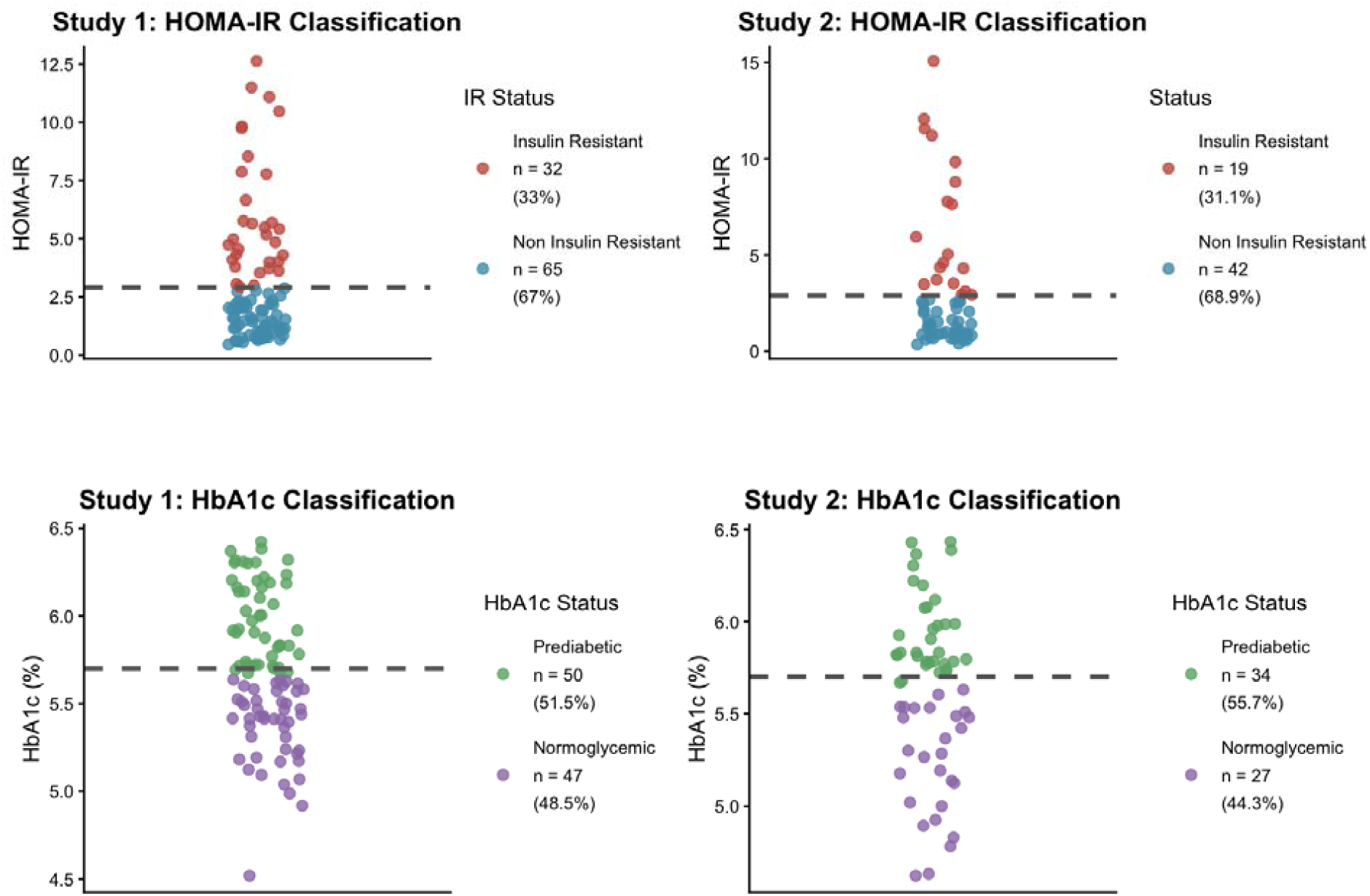
IR classification and glycemic status across Study 1 and Study 2. *Note*. Top row: participants classified by HOMA-IR using a threshold of ≥ 2.9 in Study 1 (left) and Study 2 (right). Bottom row: glycemic status classified by HbA1c of the same participants in Study 1 (left) and Study 2 (right), with Normoglycemic defined as HbA1c < 5.7%, and Prediabetic as 5.7-6.4%, per American Diabetes Association criteria. Each dot represents one participant. Dashed lines indicate clinical classification thresholds. Sample sizes and group proportions are displayed in the legends.

**Figure S3.**
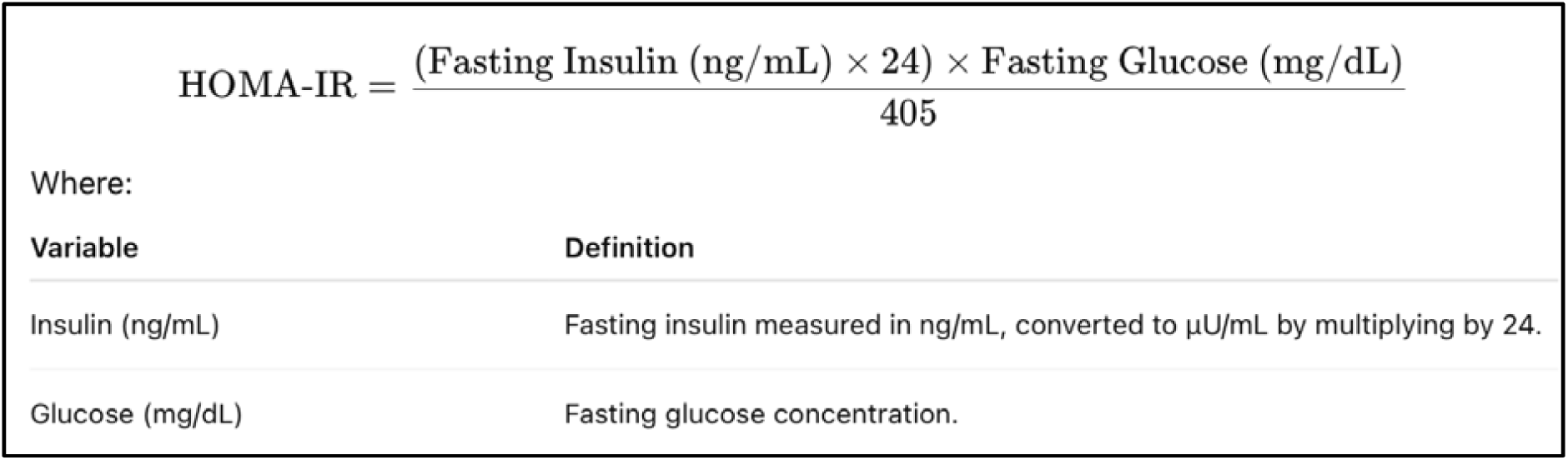
The Homeostatic Model Assessment for Insulin Resistance (HOMA-IR) equation. *Note*. *Note*. HOMA-IR is derived from the product of fasting plasma insulin (µU/mL) and fasting plasma glucose (mg/dL), divided by the constant 405 — the mg/dL-equivalent normalizing factor corresponding to the expected product of typical fasting values. Higher HOMA-IR values reflect greater insulin resistance. Equation is adapted from Ref.^18^ in the main manuscript.

**Figure S4.**
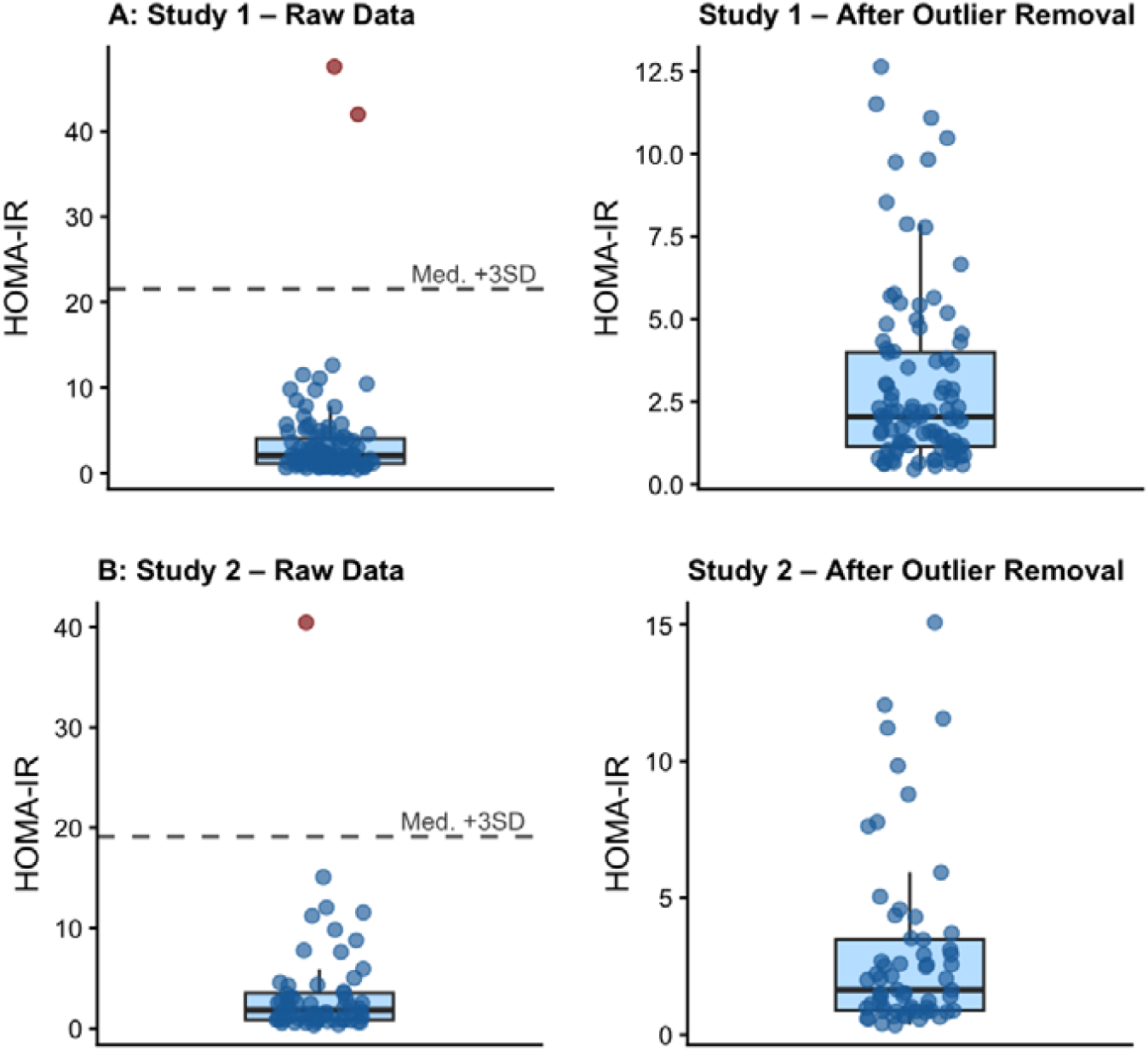
HOMA-IR distributions before and after outlier removal in Study 1 and Study 2. *Note*. Panel A shows Study 1 *(N* = 99 before exclusion) and Panel B shows Study 2 (*N* = 62 before exclusion). Left panels display raw HOMA-IR values, with outliers highlighted in red (Study 1: n = 2, HOMA-IR = 41.9 and 45.6; Study 2: n = 1, HOMA-IR = 40.4); these values exceeded the sample median by more than 3 *SD* and were excluded as failing to meet the required 8-hour fasting protocol. Right panels display the cleaned distributions after outlier removal (Study 1: n = 97; Study 2: n = 61), with the dashed horizontal line indicating the HOMA-IR binarization threshold of 2.9 used to classify participants as IR or Non-IR. Box plots display the median, interquartile range, and 1.5× IQR whiskers; individual data points are shown as jittered dots.

**Figure S5.**
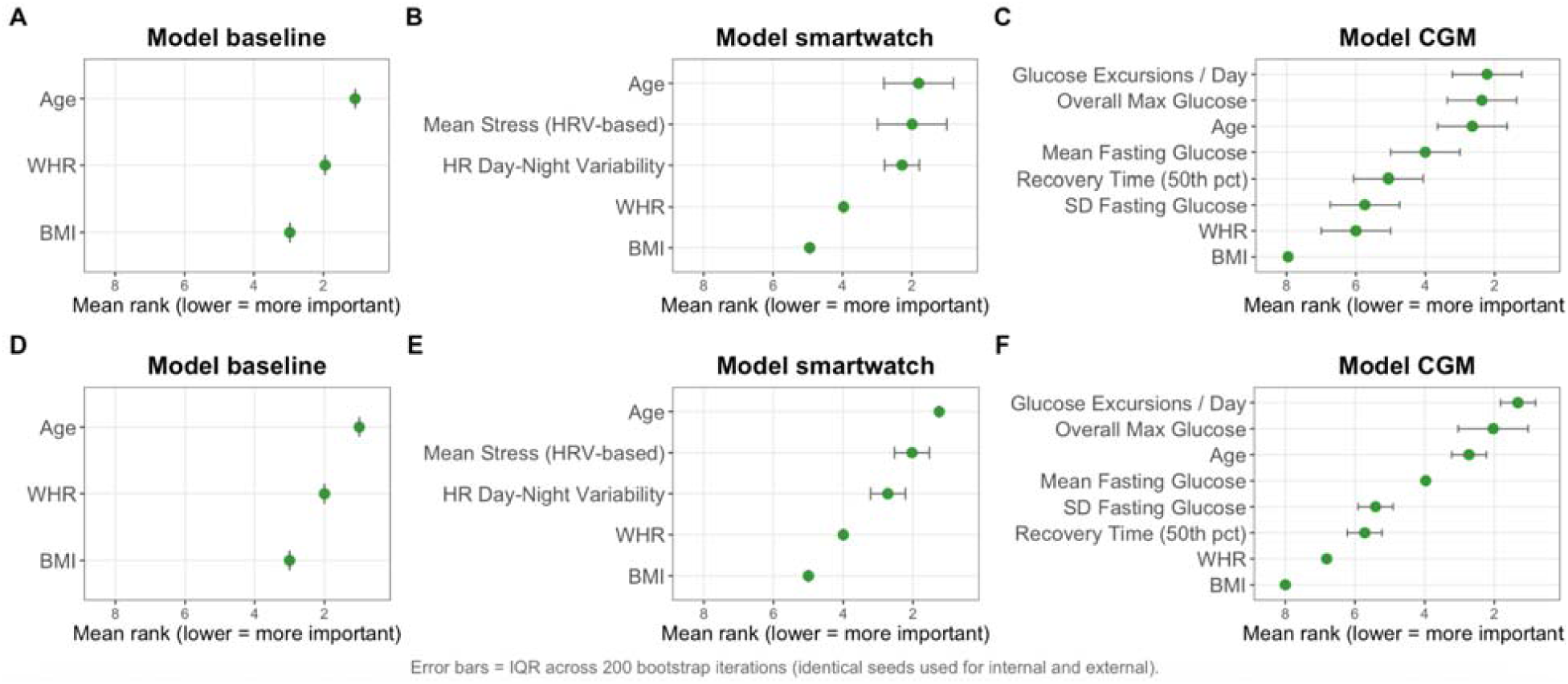
Feature-rank stability across studies *Note*. (A–C) Internal stability (Study 1). Feature-rank stability quantifies how consistently each model orders its predictors under repeated resampling. Across 200 stratified 70/30 splits of Study 1, each model is trained on ∼68 participants and evaluated on the held-out ∼29. For every split, features are ranked by their mean absolute SHAP values (rank 1 = least important; rank p = most important). Points display the mean rank across iterations, and error bars show the interquartile range (IQR), reflecting variability arising from changes to both the training set and the internal test set. Panels A–C correspond to the baseline, smartwatch, and CGM models, respectively. (D–F) External stability (Study 2). For external assessment, the same 200 seeds used in Study 1 are reused to generate 200 bootstrap resamples of the Study 2 cohort (sampled with replacement while preserving IR prevalence). Each model is retrained once on all 97 participants from Study 1, and feature ranks are recalculated for each external bootstrap sample. Points represent the mean external feature rank, and error bars represent the IQR, indicating how sensitive the feature-importance ordering is to which external participants are included in each resample. Panels D–F correspond to the baseline, smartwatch, and CGM models evaluated on Study 2.

**Table S1.** Overview of Features for the Baseline, CGM, and Smartwatch Models.

| Category | Feature | Baseline | CGM Model | Smartwatch Model |
| --- | --- | --- | --- | --- |
| **Demographic** |  |  |  |  |
|  | **Age.** Participant age (years) | ✓ | ✓ | ✓ |
| **Anthropometric** |  |  |  |  |
|  | **BMI.** Body mass index (kg/m<sup>2</sup>) | ✓ | ✓ | ✓ |
|  | **WHR.** Waist-to-hip ratio (waist circumference ÷ hip circumference) | ✓ | ✓ | ✓ |
| **Glycemic (CGM-derived)** |  |  |  |  |
|  | **Mean Fasting Glucose.** Daily mean glucose from wake to first meal (mmol/L) | — | ✓ | — |
|  | **SD Fasting Glucose.** Day-to-day variability of wake-to-first-meal glucose (mmol/L) | — | ✓ | — |
|  | **Glucose Excursions Per Day.** Mean number of glucose excursions above threshold per day | — | ✓ | — |
|  | **Recovery Time (50th percentile).** Median time for glucose to recover 50% from peak to nadir (min) | — | ✓ | — |
|  | **Overall Maximum Glucose.** Highest recorded glucose value across the monitoring period (mmol/L) | — | ✓ | — |
| **HR/ Heart Rate Variability** |  |  |  |  |
|  | **HR Day–Night Variability.** SD of the difference between daytime and nocturnal heart rate (bpm) | — | — | ✓ |
| **Stress** |  |  |  |  |
|  | **Mean Stress (HRV-based).** HRV-derived mean daily stress score from smartwatch sensor data | — | — | ✓ |
|  |  | **Baseline** | **CGM** | **Smartwatch** |
|  | No. of features | 3 | 8 | 5 |
|  | Feature domains | Anthropometric /Demographic | Anthropometric/Demographic, Glycemic | Anthropometric/Demographic, HR/HRV |
|  | mtry (floor√p) | 1 | 2 | 2 |

**Note.** ✓ = feature included; — = feature not included. *BMI* = body mass index; *WHR* = waist-to-hip ratio; *CGM* = continuous glucose monitor; *HRV* = heart rate variability; *HR* = heart rate; *SD* = standard deviation; *mtry* = number of features randomly sampled at each tree split (floor√p, where p = number of features).

**Table S2.** Descriptives of key study variables by IR status across studies.

| Variable | Study 1 (N = 97) |  |  | Study 2 (N = 61) |  |  |
| --- | --- | --- | --- | --- | --- | --- |
|  | Overall<br>(n = 97) | Non-IR<br>(n = 65) | IR<br>(n = 32) | Overall<br>(n = 61) | Non-IR<br>(n = 42) | IR<br>(n = 19) |
|  | M (SD) | M (SD) | M (SD) | M (SD) | M (SD) | M (SD) |
| **Demographic** |  |  |  |  |  |  |
| Age (years) | 57.91 (11.07) | 58.11 (10.81) | 57.50 (11.74) | 56.02 (10.24) | 56.45 (11.01) | 55.05 (8.49) |
| HOMA-IR | 2.98 (2.71) | 1.49 (0.66) | 6.02 (2.76) | 3.01 (3.29) | 1.33 (0.70) | 6.73 (3.72) |
| **Anthropometric** |  |  |  |  |  |  |
| BMI (kg/m<sup>2</sup>) | 30.23 (7.68) | 27.10 (4.63) | 36.58 (8.73) | 33.23 (9.39) | 31.03 (8.76) | 38.10 (9.09) |
| Waist-to-Hip Ratio | 0.91 (0.09) | 0.89 (0.09) | 0.95 (0.08) | 0.89 (0.09) | 0.87 (0.08) | 0.94 (0.08) |
| **Glycaemic (CGM)** |  |  |  |  |  |  |
| Mean Fasting Glucose (mg/dL) | 116.24 (15.28) | 113.14 (12.90) | 123.21 (17.98) | 114.12 (17.67) | 110.79 (15.15) | 121.15 (20.79) |
| SD Fasting Glucose (mg/dL) | 7.67 (4.37) | 6.81 (4.04) | 9.68 (4.54) | 8.89 (4.31) | 8.08 (3.99) | 10.79 (4.56) |
| Excursions per Day | 6.30 (0.70) | 6.29 (0.76) | 6.32 (0.55) | 6.55 (0.79) | 6.60 (0.80) | 6.44 (0.77) |
| Recovery Time 50% Median | 29.47 (10.31) | 28.48 (10.98) | 31.58 (8.50) | 28.57 (11.37) | 27.32 (10.99) | 31.32 (12.00) |
| Max Peak Glucose (mg/dL) | 227.80 (44.14) | 224.80 (43.58) | 234.10 (45.37) | 232.25 (53.18) | 228.17 (56.37) | 241.26 (45.42) |
| **Smartwatch HR/HRV** |  |  |  |  |  |  |
| HR Day–Night Variability | 5.66 (2.82) | 5.58 (3.05) | 5.83 (2.31) | 6.54 (2.85) | 6.40 (3.09) | 6.85 (2.27) |
| Mean Daily Stress Score | 22.33 (13.19) | 21.62 (12.39) | 23.73 (14.75) | 27.46 (13.32) | 25.11 (11.97) | 32.65 (14.95) |

*Note.* Values presented as M (SD) = mean (standard deviation). IR = insulin resistance; Non-IR = no insulin resistance; BMI = body mass index; WHR = waist-to-hip ratio; CGM = continuous glucose monitor; HR = heart rate; HRV = heart rate variability; bpm = beats per minute.

**Table S3.** HbA1c-based prediabetes and normoglycemia classification by IR Status across studies.

|  | Study 1 (N = 97) |  | Study 2 (N = 61) |  |
| --- | --- | --- | --- | --- |
|  | Non-IR<br>n (%) | IR<br>n (%) | Non-IR<br>n (%) | IR<br>n (%) |
| **HbA1c classification** |  |  |  |  |
| Normoglycemic | 38 (58.5%) | 9 (28.1%) | 23 (54.8%) | 4 (21.1%) |
| Prediabetic | 27 (41.5%) | 23 (71.9%) | 19 (45.2%) | 15 (78.9%) |
| $\chi^2(df)$ | 6.73 (1) | | 4.74 (1) | |
| *p* | .009 |  | .030 |  |
| Cramér's *V* | .26 |  | .28 |  |
| 95% CI [*V*] | [.08, .45] |  | [.06, .49] |  |

*Note.* IR = insulin resistant (HOMA-IR ≥ 2.9). HbA1c thresholds: normoglycemia < 5.7%, prediabetes 5.7–6.4%. Cramér's *V* effect sizes: small = .10, medium = .30, large = .50 (Cohen, 1988). 95% CIs for Cramér's *V* estimated via bootstrapping (2,000 resamples).

**Table S4.**
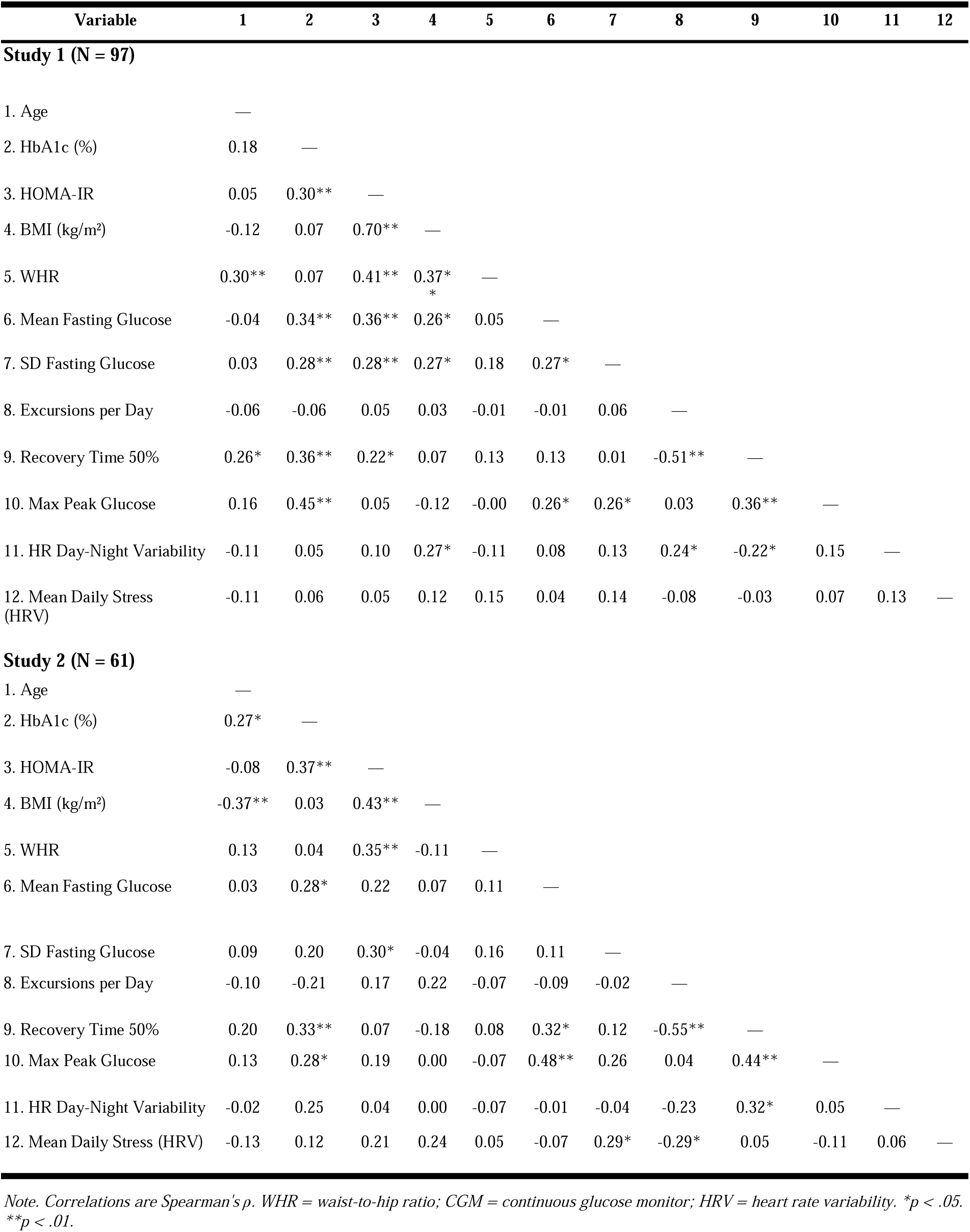
Spearman correlations among key variables across studies.

**Table S5.**
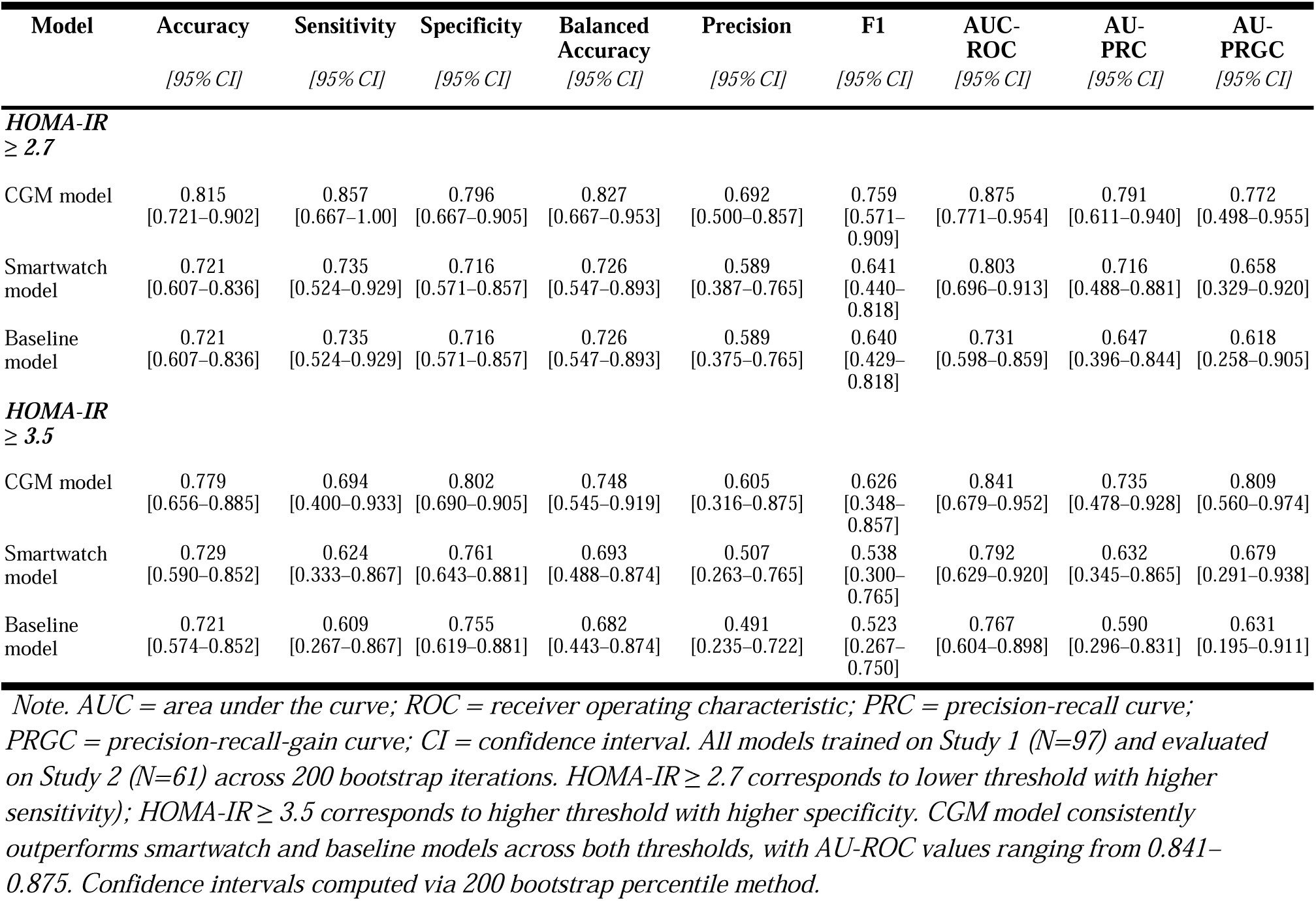
External validation (Study 2) model performance alternative HOMA-IR cut-off thresholds.

**Table S6.** External Validation model performance cross alternative imputation strategies.

| Model | Accuracy | Sensitivity | Specificity | Balanced Accuracy | Precision | F1 | AUC-ROC | AU-PRC | AU-PRGC |
| --- | --- | --- | --- | --- | --- | --- | --- | --- | --- |
|  | [95% CI] | [95% CI] | [95% CI] | [95% CI] | [95% CI] | [95% CI] | [95% CI] | [95% CI] | [95% CI] |
| **Imputation:** |  |  |  |  |  |  |  |  |  |
| **KNN-3** |  |  |  |  |  |  |  |  |  |
| CGM model | 0.795<br>[0.689–0.902] | 0.835<br>[0.619–1.000] | 0.778<br>[0.643–0.905] | 0.806<br>[0.631–0.953] | 0.688<br>[0.481–0.867] | 0.718<br>[0.522–0.875] | 0.865<br>[0.760–0.953] | 0.813<br>[0.639–0.950] | 0.872<br>[0.684–0.975] |
| Smartwatch model | 0.713<br>[0.590–0.820] | 0.730<br>[0.500–0.929] | 0.706<br>[0.564–0.843] | 0.718<br>[0.532–0.886] | 0.575<br>[0.368–0.756] | 0.614<br>[0.408–0.789] | 0.781<br>[0.649–0.893] | 0.692<br>[0.495–0.863] | 0.677<br>[0.332–0.928] |
| Baseline model | 0.721<br>[0.607–0.836] | 0.737<br>[0.529–0.929] | 0.714<br>[0.574–0.848] | 0.726<br>[0.551–0.889] | 0.583<br>[0.375–0.769] | 0.622<br>[0.429–0.800] | 0.747<br>[0.623–0.867] | 0.612<br>[0.387–0.839] | 0.615<br>[0.321–0.892] |
| **Imputation:** |  |  |  |  |  |  |  |  |  |
| **KNN-5** |  |  |  |  |  |  |  |  |  |
| CGM model | 0.800<br>[0.697–0.902] | 0.839<br>[0.625–1.000] | 0.782<br>[0.648–0.905] | 0.810<br>[0.637–0.953] | 0.692<br>[0.488–0.871] | 0.722<br>[0.528–0.882] | 0.870<br>[0.765–0.958] | 0.818<br>[0.644–0.954] | 0.876<br>[0.691–0.978] |
| Smartwatch model | 0.713<br>[0.590–0.820] | 0.730<br>[0.500–0.929] | 0.706<br>[0.564–0.843] | 0.718<br>[0.532–0.886] | 0.575<br>[0.368–0.756] | 0.614<br>[0.408–0.789] | 0.781<br>[0.649–0.893] | 0.692<br>[0.495–0.863] | 0.677<br>[0.332–0.928] |
| Baseline model | 0.721<br>[0.607–0.836] | 0.737<br>[0.529–0.929] | 0.714<br>[0.574–0.848] | 0.726<br>[0.551–0.889] | 0.583<br>[0.375–0.769] | 0.622<br>[0.429–0.800] | 0.747<br>[0.623–0.867] | 0.612<br>[0.387–0.839] | 0.615<br>[0.321–0.892] |
| **Imputation:** |  |  |  |  |  |  |  |  |  |
| **KNN-7** |  |  |  |  |  |  |  |  |  |
| CGM model | 0.805<br>[0.705–0.902] | 0.844<br>[0.643–1.000] | 0.788<br>[0.655–0.905] | 0.816<br>[0.649–0.953] | 0.698<br>[0.500–0.875] | 0.729<br>[0.545–0.887] | 0.875<br>[0.770–0.961] | 0.823<br>[0.650–0.958] | 0.882<br>[0.701–0.981] |
| Smartwatch model | 0.719<br>[0.600–0.831] | 0.736<br>[0.520–0.933] | 0.712<br>[0.571–0.848] | 0.724<br>[0.545–0.891] | 0.581<br>[0.378–0.763] | 0.620<br>[0.420–0.795] | 0.786<br>[0.654–0.898] | 0.697<br>[0.503–0.867] | 0.681<br>[0.338–0.932] |
| Baseline model | 0.721<br>[0.607–0.836] | 0.737<br>[0.529–0.929] | 0.714<br>[0.574–0.848] | 0.726<br>[0.551–0.889] | 0.583<br>[0.375–0.769] | 0.622<br>[0.429–0.800] | 0.747<br>[0.623–0.867] | 0.612<br>[0.387–0.839] | 0.615<br>[0.321–0.892] |

*Note. AUC = area under the curve; ROC = receiver operating characteristic; PRC = precision-recall curve; PRGC = precision-recall-gain curve; CI = confidence interval; KNN = k-nearest neighbors. All models trained on Study 1 (N=97) and evaluated on Study 2 (N=61) across 200 bootstrap iterations. Three imputation strategies tested: KNN-3 (k=3 neighbors), KNN-5 (k=5 neighbors), and KNN-7 (k=7 neighbors). Results demonstrate robustness of model ranking (CGM > Smartwatch > Baseline) across all imputation methods. CGM model maintains AUC-ROC $\geq 0.865$ regardless of imputation strategy (range: 0.865–0.875). Baseline model shows no variation across strategies (no missing data in baseline features). Smartwatch model shows minimal variation (AUC-ROC range: 0.781–0.786). Confidence intervals computed via bootstrap percentile method.*

### Supplement A: Differences among study samples

The two study samples were largely comparable across demographic and clinical characteristics. Age did not differ significantly between studies (Study 1: *M* = 57.9, Median = 57.0; Study 2: *M* = 56.0, Median = 56.0; *U* = 3233.5, *p* = .327), nor did the proportion classified as insulin resistant (33.0% vs. 31.1%; χ²(1) = 0.004, *p* = .947). HbA1c was similar across studies (Study 1: *M* = 5.69%, Median = 5.70; Study 2: *M* = 5.63%, Median = 5.70; *U* = 3053.5, *p* = .735). When dichotomized at the clinical threshold (HbA1c ≥5.7%), Study 2 showed a directionally higher prevalence of prediabetes (55.7%, *n*=34) compared to Study 1 (51.5%, *n*=50), though this difference was not statistically significant (χ²(1) = 0.123, *p* = .726). Fasting insulin was comparable (Study 1: *M* = 88.1 pmol/L, Median = 62.0; Study 2: *M* = 97.3 pmol/L, Median = 67.2; *U* = 2905.5, *p* = .851), as was waist-to-hip ratio (Study 1: *M* = 0.91, Median = 0.91; Study 2: *M* = 0.89, Median = 0.90; *U* = 3280.0, *p* = .277), and CGM wear days (Study 1: *M* = 10.8, Median = 11.0; Study 2: *M* = 10.7, Median = 11.0; *U* = 3045.0, *p* = .424). However, fasting glucose was significantly higher in Study 1 than Study 2 (*M* = 5.32 vs. 4.64 mmol/L; *U* = 4445.5, *p* < .001). Study 2 participants had significantly higher BMI (*M* = 33.2 vs. 30.2 kg/m²; *U* = 2316.0, *p* = .022) and lower number of smartwatch days (*M* = 18.4 Study 1 vs. 15.2 days Study 2; *U* = 3789.0, *p* = .003).

Most CGM features were comparable across studies. Mean fasting glucose did not differ significantly (*M* = 116.4 vs. 113.5 mg/dL; *U* = 2820.0, *p* = .185), nor did max peak glucose (*M* = 227.8 vs. 232.3 mg/dL; *U* = 2928.0, *p* = 1.00) or PPG recovery rate (*M* = 0.84 vs. 0.72; *U* = 2701.0, *p* = .544). However, SD fasting glucose — the day-to-day variability of wake-to-meal glucose — was significantly higher in Study 2 (*M* = 8.75 vs. 7.41 mg/dL; *U* = 1694.0, *p* = .018), suggesting greater within-person glycemic instability in the external sample. Both smartwatch-derived features differed significantly between studies. HR day–night variability was higher in Study 2 (*M* = 6.54 vs. 5.66; *U* = 2092.0, *p* = .029), and mean daily stress score was also significantly higher in Study 2 (*M* = 27.5 vs. 22.3; *U* = 2246.0, *p* = .014).

### Supplement B: Physical Activity Analyses

To ensure a comprehensive evaluation of lifestyle wearable–derived data modalities, we conducted a feature search across common physical activity (PA) features derived from the Garmin Vivosmart 5. PA features included daily mean and variability of step count, active minutes, step distribution across morning, afternoon, and evening windows, activity onset and offset timing, intra-daily variability, and inter-daily stability, averaged over the course of the observational period (approximately ten days), and accounting for ten features in total.

External validation results from Study 2 showed that PA features alone performed poorly (AU-ROC = 0.511 [0.380–0.678]), worse than random guessing. Adding PA features to the baseline (anthropometrics +age) model did not improve performance: the baseline model achieved AU-ROC = 0.749 [0.601–0.868], nearly identical to the PA + Baseline model at 0.753 [0.609–0.877]. A similar pattern was observed for the smartwatch model, where AU-ROC = 0.789 [0.647–0.904] without PA and 0.788 [0.646–0.894] with PA, indicating no gain. For the CGM model, which performed strongest (AU-ROC = 0.873 [0.756–0.967]), adding PA also produced no improvement (PA + CGM AU-ROC = 0.851 [0.740–0.961]). The combined multimodal model excluding PA features (CGM + Smartwatch + Baseline) (AU-ROC = 0.865 [0.740–0.954]) and exceeded the performance of the model including all features (AU-ROC = 0.853 [0.715–0.955]).

The combined multimodal model excluding PA features (CGM + Smartwatch + Baseline) (AU-ROC = 0.865 [0.740–0.954]) and exceeded the performance of the model including all features (AU-ROC = 0.853 [0.715–0.955]). Overall, the commonly extracted PA features derived from the Garmin Vivosmart 5 did not contribute meaningful independent predictive signals for IR in this cohort, whether used alone or added to anthropometric, smartwatch-based HR/HRV, or CGM-derived features.

**Table S7.** Physical activity feature Evaluation: external validation performance (Study 2)

| **Model** | **AU-ROC**<br>**[95% CI]** | **AU-PRC**<br>**[95% CI]** | **Accuracy** | **Sensitivity** | **Specificity** | **Balanced**<br>**Accuracy** | **Precision** | **F1** |
| --- | --- | --- | --- | --- | --- | --- | --- | --- |
| PA alone | 0.511<br>[0.380–0.678] | 0.344<br>[0.251–0.522] | 0.557 | 0.211 | 0.714 | 0.462 | 0.250 | 0.229 |
| PA + Baseline | 0.753<br>[0.609–0.877] | 0.585<br>[0.404–0.873] | 0.770 | 0.632 | 0.833 | 0.732 | 0.632 | 0.632 |
| PA + Smartwatch | 0.788<br>[0.646–0.894] | 0.635<br>[0.433–0.887] | 0.787 | 0.632 | 0.857 | 0.744 | 0.667 | 0.649 |
| PA + CGM | 0.851<br>[0.740–0.961] | 0.769<br>[0.638–0.935] | 0.820 | 0.632 | 0.905 | 0.768 | 0.750 | 0.686 |
| CGM + Smartwatch +<br>Baseline | 0.865<br>[0.740–0.954] | 0.780<br>[0.624–0.907] | 0.869 | 0.737 | 0.929 | 0.833 | 0.824 | 0.778 |
| All features (inc. PA) | 0.853<br>[0.715–0.955] | 0.760<br>[0.569–0.917] | 0.820 | 0.632 | 0.905 | 0.768 | 0.750 | 0.686 |

*Note. AU-ROC = area under the receiver operating characteristic curve; AU-PRC = area under the precision-recall curve; CI = confidence interval. All models trained on Study 1 (N=97) and evaluated on Study 2 (N=61) using 50 bootstrap iterations, ntree=300, median imputation, stratified upsampling, mtry=floor( $\sqrt{p}$ ). PA features include: daily mean and variability of step count, active minutes, step distribution across morning/afternoon/evening, activity onset and offset timing, and intra-daily variability (10 features total). Results demonstrate that PA features alone (AU-ROC = 0.511) perform worse than random guessing. Adding PA features to baseline (0.755 vs 0.753), smartwatch (0.792 vs 0.788), or CGM (0.861 vs 0.851) models shows no improvement or marginal degradation. The combined model without PA (CGM + Smartwatch + Baseline: 0.865) outperforms the model with all features including PA (0.853), confirming that PA features do not contribute independent predictive signals for IR in the given cohort.*

## References

1. World Health Organization. Diabetes: Fact sheet. World Health Organization. Available at: https://www.who.int/news-room/fact-sheets/detail/diabetes. Accessed 2026.

2. International Diabetes Federation. IDF Diabetes Atlas, 11th edn. Brussels: International Diabetes Federation (2025). Available at: https://diabetesatlas.org.

3. NCD Risk Factor Collaboration (NCD-RisC). Worldwide trends in diabetes prevalence and treatment from 1990 to 2022: a pooled analysis of 1108 population-representative studies with 141 million participants. Lancet 404, 2077–2093 (2024).

4. Diabetes Prevention Program Research Group. Reduction in the Incidence of Type 2 Diabetes with Lifestyle Intervention or Metformin. (2002) doi:10.1056/NEJMoa012512.

5. Tuomilehto, J. et al. Prevention of type 2 diabetes mellitus by changes in lifestyle among subjects with impaired glucose tolerance. The New England journal of medicine 344, (2001).

6. Roden, M. & Shulman, G. I. The integrative biology of type 2 diabetes. Nature 576, 51–60 (2019).

7. Chen, S. et al. The global macroeconomic burden of diabetes mellitus. Nat Med 32, 126–138 (2026).

8. Davies, M. J. et al. Type 2 diabetes mellitus. Nat Rev Dis Primers 12, (2026).

9. DeFronzo, R. A. et al. Type 2 diabetes mellitus. Nat Rev Dis Primers 1, 15019 (2015).

10. Weyer, C., Bogardus, C., Mott, D. M. & Pratley, R. E. The natural history of insulin secretory dysfunction and insulin resistance in the pathogenesis of type 2 diabetes mellitus. The Journal of clinical investigation 104, (1999).

11. Tabák, A. G. et al. Trajectories of glycaemia, insulin sensitivity, and insulin secretion before diagnosis of type 2 diabetes: an analysis from the Whitehall II study. Lancet 373, 2215–2221 (2009).

12. Tabák, A. G., Herder, C., Rathmann, W., Brunner, E. J. & Kivimäki, M. Prediabetes: a high-risk state for diabetes development. Lancet 379, 2279–2290 (2012).

13. Færch, K. et al. Trajectories of cardiometabolic risk factors before diagnosis of three subtypes of type 2 diabetes: a post-hoc analysis of the longitudinal Whitehall II cohort study. Lancet Diabetes Endocrinol 1, 43–51 (2013).

14. Uusitupa, M. et al. Long-term improvement in insulin sensitivity by changing lifestyles of people with impaired glucose tolerance: 4-year results from the Finnish Diabetes Prevention Study. Diabetes 52, (2003).

15. Wang, Y. et al. Effectiveness of Different Intervention Modes in Lifestyle Intervention for the Prevention of Type 2 Diabetes and the Reversion to Normoglycemia in Adults With Prediabetes: Systematic Review and Meta-Analysis of Randomized Controlled Trials. J Med Internet Res 27, e63975 (2025).

16. Pan, X. R. et al. Effects of diet and exercise in preventing NIDDM in people with impaired glucose tolerance. The Da Qing IGT and Diabetes Study. Diabetes Care 20, 537–544 (1997).

17. Muniyappa, R., Lee, S., Chen, H. & Quon, M. J. Current approaches for assessing insulin sensitivity and resistance in vivo: advantages, limitations, and appropriate usage. Am J Physiol Endocrinol Metab 294, E15–26 (2008).

18. Matthews, D. R. et al. Homeostasis model assessment: insulin resistance and beta-cell function from fasting plasma glucose and insulin concentrations in man. Diabetologia 28, 412–419 (1985).

19. Bonora, E. et al. Homeostasis model assessment closely mirrors the glucose clamp technique in the assessment of insulin sensitivity: studies in subjects with various degrees of glucose tolerance and insulin sensitivity. Diabetes Care 23, 57–63 (2000).

20. Duncan, B. B., Magliano, D. J. & Boyko, E. J. IDF Diabetes Atlas 11th edition 2025: global prevalence and projections for 2050. Nephrol Dial Transplant 41, 7–9 (2025).

21. Website. https://www.statista.com/outlook/hmo/digital-health/digital-fitness-well-being/fitness-trackers/smartwatches/worldwide.

22. Kumar, N. Smartwatch Statistics (2026): Global Users & Market Share. DemandSage https://www.demandsage.com/smartwatch-statistics/ (2026).

23. Website. https://www.fda.gov/news-events/press-announcements/fda-clears-first-over-counter-continuous-glucose-monitor.

24. Liao, X. et al. Continuous glucose monitoring in non-diabetic populations: a systematic review of observational and interventional studies with meta-analysis. European Journal of Medical Research 31, 397 (2026).

25. Wilczek, F., van der Stouwe, J. G., Petrasch, G. & Niederseer, D. Non-Invasive Continuous Glucose Monitoring in Patients Without Diabetes: Use in Cardiovascular Prevention-A Systematic Review. Sensors (Basel) 25, (2025).

26. Svensson, M. K. et al. Alterations in heart rate variability during everyday life are linked to insulin resistance. A role of dominating sympathetic over parasympathetic nerve activity? Cardiovasc Diabetol 15, 91 (2016).

27. Saito, I. et al. Heart Rate Variability, Insulin Resistance, and Insulin Sensitivity in Japanese Adults: The Toon Health Study. J Epidemiol 25, 583–591 (2015).

28. Hajj-Boutros, G., Landry-Duval, M.-A., Comtois, A. S., Gouspillou, G. & Karelis, A. D. Wrist-worn devices for the measurement of heart rate and energy expenditure: A validation study for the Apple Watch 6, Polar Vantage V and Fitbit Sense. Eur J Sport Sci 23, 165–177 (2023).

29. Poon, A. K. et al. Insulin resistance and reduced cardiac autonomic function in older adults: the Atherosclerosis Risk in Communities study. BMC Cardiovasc Disord 20, 217 (2020).

30. Perciaccante, A., Fiorentini, A., Paris, A., Serra, P. & Tubani, L. Circadian rhythm of the autonomic nervous system in insulin resistant subjects with normoglycemia, impaired fasting glycemia, impaired glucose tolerance, type 2 diabetes mellitus. BMC Cardiovasc Disord 6, 19 (2006).

31. Metwally, A. A. et al. Prediction of metabolic subphenotypes of type 2 diabetes via continuous glucose monitoring and machine learning. Nature Biomedical Engineering 9, 1222–1239 (2024).

32. Hall, H. et al. Glucotypes reveal new patterns of glucose dysregulation. PLoS Biol 16, e2005143 (2018).

33. Metwally, A. A. et al. Insulin resistance prediction from wearables and routine blood biomarkers. Nature 652, 451–461 (2026).

34. Cutillo, C. M. et al. Machine intelligence in healthcare—perspectives on trustworthiness, explainability, usability, and transparency. npj Digital Medicine 3, 47 (2020).

35. Owsley, C. et al. Cross-sectional design and protocol for Artificial Intelligence Ready and Equitable Atlas for Diabetes Insights (AI-READI). BMJ Open 15, e097449 (2025).

36. AI-READI Consortium. AI-READI: rethinking AI data collection, preparation and sharing in diabetes research and beyond. Nat Metab 6, 2210–2212 (2024).

37. Fuchs, M. et al. Predicting individual differences in digital alcohol intervention effectiveness through multimodal data. NPJ Digit Med 9, 170 (2026).

38. Brügger, V., Kowatsch, T. & Jovanova, M. Predicting postprandial glucose excursions to personalize dietary interventions for type-2 diabetes management. Scientific Reports 15, 25920 (2025).

39. Rose, S. Machine Learning for Prediction in Electronic Health Data. JAMA network open 1, (2018).

40. Flach, P. & Kull, M. Precision-Recall-Gain Curves: PR Analysis Done Right. Advances in Neural Information Processing Systems 28, (2015).

41. Artificial Intelligence Ready and Exploratory Atlas for Diabetes Insights. AI-READI https://aireadi.org/.

42. AI-READI Consortium. Flagship Dataset of Type 2 Diabetes from the AI-READI Project. FAIRhub 10.34534/2 (2024).

43. Liu, J., Jin, X., Feng, Z. & Huang, J. Using anthropometric parameters to predict insulin resistance among patients without diabetes mellitus. Scientific Reports 14, 21407 (2024).

44. Jamar, G. et al. Evaluation of waist-to-height ratio as a predictor of insulin resistance in non-diabetic obese individuals. A cross-sectional study. Sao Paulo Med. J. 135, 462–468 (2017).

45. Steyerberg, E. W. Clinical Prediction Models. (Springer New York).

46. Nohara, Y., Matsumoto, K., Soejima, H. & Nakashima, N. Explanation of machine learning models using shapley additive explanation and application for real data in hospital. Computer methods and programs in biomedicine 214, 106584 (2022).

47. Van Calster, B. et al. Reporting and Interpreting Decision Curve Analysis: A Guide for Investigators. Eur Urol 74, 796–804 (2018).

48. Ascaso, J. F. et al. Diagnosing insulin resistance by simple quantitative methods in subjects with normal glucose metabolism. Diabetes care 26, (2003).

49. Ballena-Caicedo, J. et al. Global prevalence of insulin resistance in the adult population: a systematic review and meta-analysis. Front. Endocrinol. 16, 1646258 (2025).

50. Lebovitz, H. E. Insulin resistance--a common link between type 2 diabetes and cardiovascular disease. Diabetes Obes Metab 8, 237–249 (2006).

51. Horton, W. B., Love, K. M., Gregory, J. M., Liu, Z. & Barrett, E. J. Metabolic and vascular insulin resistance: partners in the pathogenesis of cardiovascular disease in diabetes. American Journal of Physiology-Heart and Circulatory Physiology (2025) doi:10.1152/ajpheart.00826.2024.

52. Coravos, A., Khozin, S. & Mandl, K. D. Developing and adopting safe and effective digital biomarkers to improve patient outcomes. NPJ Digit Med 2, (2019).

53. Ko, G., Davidson, L. E., Brennan, A. M., Lam, M. & Ross, R. Abdominal Adiposity, Not Cardiorespiratory Fitness, Mediates the Exercise-Induced Change in Insulin Sensitivity in Older Adults. PloS one 11, (2016).

54. Gayoso-Diz, P. et al. Insulin resistance (HOMA-IR) cut-off values and the metabolic syndrome in a general adult population: effect of gender and age: EPIRCE cross-sectional study. BMC Endocr Disord 13, 47 (2013).

55. Lee, C. H. et al. Optimal Cut-Offs of Homeostasis Model Assessment of Insulin Resistance (HOMA-IR) to Identify Dysglycemia and Type 2 Diabetes Mellitus: A 15-Year Prospective Study in Chinese. PLOS ONE 11, e0163424 (2016).

56. Mekniran, W. et al. Early health technology assessment of digital diabetes screening in Switzerland: cost-effectiveness and budget impact analyses. medRxiv 2026.02.10.26345992 (2026) doi:10.64898/2026.02.10.26345992.

57. Mekniran, W. et al. Digital health technologies and stakeholder incentives in type-2 diabetes prevention. DIGITAL HEALTH (2026) doi:10.1177/20552076261425402.

58. Giger, O.-F., Fleisch, E., Jovanova, M. & Kowatsch, T. Barriers and facilitators of implementing value-based care: The case of SwissDiabeter. Digit Health 11, 20552076251336322 (2025).

59. Giger, O.-F. et al. Digital health technologies and innovation patterns in diabetes ecosystems. Digit Health 11, 20552076241311740 (2025).

60. Gado, M., Tsaousidou, E., Bornstein, S. R. & Perakakis, N. Sex-based differences in insulin resistance. The Journal of endocrinology 261, (2024).

61. Brügger, V. et al. GLOW UP Study: Protocol for an Observational Digital Biomarker Study for Prediabetes Screening and Digital Phenotyping. medRxiv 2026.03.11.26348172 (2026) doi:10.64898/2026.03.11.26348172.

62. Lehmann, V. et al. Noninvasive Hypoglycemia Detection in People With Diabetes Using Smartwatch Data. Diabetes Care 46, 993–997 (2023).

63. Widatalla, N., Al Younis, S., Ghosh, S. K. & Khandoker, A. Symbolic heart rate transition motifs during nocturnal sleep are associated with diabetic complications in type 2 diabetes. PLoS One 20, e0333067 (2025).

64. Rosenbach, H., Itzkovitch, A., Gidron, Y. & Schonberg, T. Assessing Stress Level Scores Against Wearables-Driven Physiological Measurements. Stress Health 41, e70125 (2025).

65. Montaser, E., Fabris, C. & Kovatchev, B. Essential Continuous Glucose Monitoring Metrics: The Principal Dimensions of Glycemic Control in Diabetes. Diabetes Technol Ther 24, 797–804 (2022).

66. Shilo, S. et al. Continuous glucose monitoring and intrapersonal variability in fasting glucose. Nat Med 30, 1424–1431 (2024).

67. Bent, B. et al. Engineering digital biomarkers of interstitial glucose from noninvasive smartwatches. npj Digital Medicine 4, 89 (2021).

68. Kinny, F., Läer, S. & Obarcanin, E. Continuous Glucose Monitoring under standardised conditions regarding diet, exercise and stress in Healthy Young People (CGM-HYPE study): An exploratory clinical trial. PLOS Digit Health 4, e0001087 (2025).

69. Mao, Y. et al. Stratification of Patients with Diabetes Using Continuous Glucose Monitoring Profiles and Machine Learning. Health data science 2022, (2022).

70. Breiman, L. Random Forests. Machine Learning 45, 5–32 (2001).

71. A unified approach to interpreting model predictions. https://dl.acm.org/doi/10.5555/3295222.3295230 doi:10.5555/3295222.3295230.

72. Hur, S. et al. Comparison of SHAP and clinician friendly explanations reveals effects on clinical decision behaviour. npj Digital Medicine 8, 578 (2025).

73. Kuhn, M. Building Predictive Models in R Using the caret Package. J. Stat. Soft. 28, 1–26 (2008).

